# Health inequities in SARS-CoV-2 infection, seroprevalence, and COVID-19 vaccination: Results from the East Bay COVID-19 study

**DOI:** 10.1101/2021.12.02.21266871

**Authors:** Cameron Adams, Mary Horton, Olivia Solomon, Marcus Wong, Sean L. Wu, Sophia Fuller, Xiaorong Shao, Indro Fedrigo, Hong L. Quach, Diana L. Quach, Michelle Meas, Luis Lopez, Abigail Broughton, Anna L. Barcellos, Joan Shim, Yusef Seymens, Samantha Hernandez, Magelda Montoya, Darrell M. Johnson, Kenneth B. Beckman, Michael P. Busch, Josefina Coloma, Joseph A. Lewnard, Eva Harris, Lisa F. Barcellos

## Abstract

Comprehensive data on transmission mitigation behaviors and both SARS-CoV-2 infection and serostatus are needed from large, community-based cohorts to identify COVID-19 risk factors and the impact of public health measures. From July 2020-March 2021, approximately 5,500 adults from the East Bay Area, California were followed over three data collection rounds to investigate the association between geographic and demographic characteristics and transmission mitigation behavior with SARS-CoV-2 prevalence. We estimated the populated-adjusted prevalence of antibodies from SARS-CoV-2 infection and COVID-19 vaccination, and self-reported COVID-19 test positivity. Population-adjusted SARS-CoV-2 seroprevalence was low, increasing from 1.03% (95% CI: 0.50-1.96) in Round 1 (July-September 2020), to 1.37% (95% CI: 0.75-2.39) in Round 2 (October-December 2020), to 2.18% (95% CI: 1.48-3.17) in Round 3 (February-March 2021). Population-adjusted seroprevalence of COVID-19 vaccination was 21.64% (95% CI: 19.20-24.34) in Round 3, with Whites having 4.35% (95% CI: 0.35-8.32) higher COVID-19 vaccine seroprevalence than non-Whites. No evidence for an association between transmission mitigation behavior and seroprevalence was observed. Despite >99% of participants reporting wearing masks, non-Whites, lower-income, and lower-educated individuals had the highest SARS-CoV-2 seroprevalence and lowest vaccination seroprevalence. Results demonstrate that more effective policies are needed to address these disparities and inequities.

## Text

The first confirmed severe acute respiratory syndrome coronavirus 2 (SARS-CoV-2) infection in the California Bay Area was reported on February 28, 2020.[1] Measures to prevent transmission were implemented soon after and included shelter-in-place orders, mask mandates, business and school closures, and social distancing recommendations. Despite these measures, spikes in reported infections occurred from July to September 2020 and December 2020 to February 2021.[2]

Local public health and healthcare systems experienced major challenges in preventing infections, identifying COVID-19 cases, and ensuring adherence to transmission mitigation strategies. Furthermore, disparities and inequities in COVID-19 have been observed in the United States[3] and there are few studies investigating the association of risk behaviors with infection outcomes at the individual level.[4–6] A detailed understanding of the effectiveness of transmission mitigation behaviors and sociodemographic factors that contribute to the disproportionate impact of COVID-19 in vulnerable communities is critical. Public health and policy directives aimed at controlling ongoing transmission, developing future prevention strategies, and targeting health disparities and inequities must be evidence-based. Large population-representative cohorts with individual-level data on social and behavioral factors[7] associated with COVID-19 and SARS-CoV-2 infection and serostatus[8–12] are needed.

To address this need, we investigated individual-level characteristics and mitigation behaviors that contributed to SARS-CoV-2 seroprevalence, self-reported infection, and viral infection, and other outcomes in a large, population-based sample of over 5,500 individuals from 12 East Bay cities in Northern California followed longitudinally. During three time periods from July 2020 to April 2021, we estimated the population-adjusted prevalence of SARS-CoV-2 outcomes and differences by age, sex, race/ethnicity, ZIP code, and other demographic strata as well as the effect of transmission mitigation behavior on SARS-CoV-2 prevalence.

## Methods

### Recruitment and participants

Recruitment and selection of study participants was completed using a screening phase followed by a longitudinal study phase with three rounds of data collection. In the screening phase, all residential addresses within the East Bay cities and communities of Albany, Berkeley, El Cerrito, El Sobrante, Emeryville, Hercules, Kensington, Oakland, Piedmont, Pinole, Richmond, and San Pablo (∼307,000 residential households) were mailed an invitation to complete a consent form and screening questionnaire. In a household, the individual aged 18 years or older with the next birthday was eligible to participate. Additional eligibility criteria included living within the study region, willingness to provide biospecimens and questionnaire responses, ability to read and speak English or Spanish, and having internet access and an email address.

The target sample size was 5,500 participants. The distribution of racial and ethnic identification in the screening questionnaire responses was more White and non-Hispanic than the region population. To obtain a sample that resembled the racial and ethnic proportions in the 2018 American Community Survey (ACS) for the study region, we ranked assigned ranks for order of inclusion to all eligible individuals who responded to the screening questionnaire. Black and/or Hispanic individuals were ranked the highest followed by other non-Whites. Order of inclusion for Whites was randomly sampled. In Round 1, individuals ranked between 1 and 5,500 were offered study enrollment. Those who declined to enroll were replaced with next highest ranked individuals who had not been offered study entry. In subsequent rounds, individuals who had participated in the previous round(s) were offered participation in the next round. If participation was declined, individuals from the pool of participants who had not participated in a study round were invited. Approximate dates for each round were July-September 2020, October-December 2020, and February-March 2021.

All participants provided their informed consent for the screening phase. All participants in the study phase provided their informed consent for each study round. The study was approved by the University of California, Berkeley Committee on Protection of Human Subjects (Protocol #2020-03-13121).

### Study procedures

At the start of each round, eligible participants were invited to participate. Those who agreed to participate received a kit via FedEx containing materials for self-collection of biospecimens, pre-paid return shipping labels, and instructions to complete an online-administered questionnaire at the same time as biospecimen collection.

### Questionnaire

The questionnaire addressed sex, gender, age, race/ethnicity, income, employment, physical and mental health, as well as symptoms potentially related to COVID-19 within the previous 2 weeks, and SARS-CoV-2 testing outside of the study. Participants were also asked about transmission mitigation behaviors including physical distancing practices, close contacts with others, and mask wearing. Questionnaires were available in English or Spanish.

### SARS-CoV-2 viral and antibody testing

Anterior nasal nare swabs for viral RNA testing and dried blood spots (DBS) for antibody testing were collected from participants at each study round. Quantitative reverse transcription PCR (RT-qPCR) was used to identify SARS-CoV-2 viral infection. Three tests were used to assess anti-SARS-CoV-2 antibodies in DBS: Ortho VITROS® Anti-SARS-CoV-2 Total IgG and spike IgG ELISA targeted antibodies against the SARS-CoV-2 spike protein (indicating prior natural infection or vaccination), and Roche-NC Total IgG targeted antibodies against the nucleocapsid (NC) protein (indicating prior natural infection only).[13] Before COVID-19 vaccines were available in the study region during rounds 1 and 2, detection of antibodies against the SARS-CoV-2 spike protein was considered evidence of SARS-CoV-2 infection. During Round 3, vaccinations were widely available, therefore detection of antibodies against the NC protein were considered evidence of SARS-CoV-2 infection, while detection of antibodies against the spike protein were considered evidence of SARS-CoV-2 infection or COVID-19 vaccination (Supplement S-4.2, Figure S-1).

### SARS-CoV-2 outcomes

The following outcomes were investigated in each round: (1) cumulative SARS-CoV-2 antibody positivity, (2) participants’ self-reported history of SARS-CoV-2 positivity from RT-qPCR testing outside the study, (3) and a surveillance definition of “probable COVID-19 case” derived from self-reported symptoms and close contact with infected individuals[14], and (4) viral SARS-CoV-2 positivity from RT-qPCR testing of nasal swabs (Supplement 4.2). We also investigated antibodies induced by COVID-19 vaccination only in Round 3. SARS-CoV-2 antibody positivity was defined as having detectable antibodies in the current and/or previous round(s). DBS samples that tested negative for anti-NC antibodies and positive for anti-spike antibodies in Round 3 were considered to have antibodies from COVID-19 vaccination alone. Self-reported COVID-19 viral positivity prevalence was defined as the proportion of participants reporting a positive viral test among all participants who reported being tested within a study round. Probable COVID-19 case prevalence was defined as the proportion of participants identified as a probable COVID-19 case among all participants who provided valid responses within a study round. Viral positivity prevalence was defined as testing positive by RT-qPCR from nasal swab samples.

## Statistical analysis

### Population-adjusted seroprevalence and other SARS-CoV-2 outcomes

Bayesian multilevel regression and poststratification (MRP) was used to estimate population-adjusted cumulative seroprevalence, self-reported SARS-CoV-2 viral positivity prevalence at each study round, and “probable COVID-19” prevalence at each study round. MRP is a regression-based method for estimating population and sub-population averages from survey data that has been shown to perform better than survey weighting, particularly with sparse data [15,16].

In addition to estimation of regional prevalence of our outcomes, we estimated prevalence within demographic groups and geographic areas in the study region. Variables of interest were gender, age, race, Hispanic ethnicity, income, education, household size, and ZIP code. We used a method described by Leeman and colleagues to generate a synthetic population for poststratification using data from the 2018 American Community Survey (ACS) and the Public Use Microdata Sample.[16] Poststratification was done using binary sex because gender is not reported by the ACS. Race and ethnicity were combined into a single variable to reduce the number of poststratification strata.

At each study round, binary SARS-CoV-2 outcomes were modeled as a function of geographic and demographic characteristics using multilevel logistic regression models. Participant sex was included as a fixed effect. Vectors of random intercepts were defined for each category of race/ethnicity, age, education, income, household size, and ZIP code and two-way interactions between ZIP code, race/ethnicity, educational attainment, income, and age. To improve estimation of geographic effects we allowed for spatial correlations using the modified Besag-York-Mollié model and included the proportion of Spanish speaking households within the ZIP code of residence from the ACS as a fixed effect (Supplement S-6).[17]

### SARS-CoV-2 outcome prevalence and measures of association

We report populated-adjusted prevalence of SARS-CoV-2 outcomes across the study region and within geographic and demographic groups of interest. To calculate prevalence estimates, posterior distributions of the relevant poststratification stratum were aggregated. We also estimated prevalence differences (PD) and prevalence ratios (PR) for the association between populated-adjusted SARS-CoV-2 outcomes and race/ethnicity, education, and sex. For each parameter of interest, the mean of the posterior distribution was the point estimate, and the 95% credible interval (CI) was the 2.5% and 97.5% quantiles of a posterior distribution.

### SARS-CoV-2 test-kit bias corrected seroprevalence

We estimated cumulative SARS-CoV-2 seroprevalence at each study round adjusted for the net sensitivity and specificity of the SARS-CoV-2 antibody testing algorithm in each study round (Supplement S-6).

### Transmission mitigation behavior analyses

In each round, participants were asked about physical distancing practices, recent close contacts with others, mask wearing, and other behaviors and activities that might affect the risk of SARS-CoV-2 infection. We classified participants into two behavior categories, “high-risk” and “low-risk”, with those responses using latent class analysis.[18] Crude associations between behavior categories and characteristics such as sex, age, race/ethnicity, education, and income were assessed with χ^2^ tests. Associations between high-risk vs. low-risk behavior and within round SARS-CoV-2 seroprevalence and self-reported test positivity were estimated using the MRP model described above with random intercepts for behavior categories and interactions between the behavior categories and ZIP code, age, race/ethnicity, education, and income (Supplement S-5 and S-6).

Statistical analyses were completed in R 4.0.2. NIMBLE was used to implement MRP models.[19] Detailed descriptions of MRP methods and code are provided in Supplementary Methods and at github.com/adams-cam/ebcovid_prev.

## Results

### Enrollment and characteristics of study participants

Of the 16,115 residents who consented and completed the screening procedures between May-July 2020, 1,777 did not satisfy inclusion criteria and were excluded (Fig 1). Characteristics of participants are presented in Table 1 and Table S-1. Participation rates were high (Round 1: 76.8%, Round 2: 89.8%, and Round 3: 87.3%), and participants identified predominantly as female (∼63%). Those aged 45-64 years were the largest age group of participants across all study rounds (ranging from 37.3% to 39.4%). Most participants identified as White (52.5% to 63.3%), followed by Asian/Pacific Islander (13.9% to 15.7%), Hispanic (11.0% to 15.6%), two or more races (6.9% to 9.1%), African American or Black (3.0% to 4.9%), and Native American/Alaska Native or other (1.7% to 2.2%).

**Table 1.** Characteristics of participants at each round of the study compared to study region population.

|  | PS Strata<sup>a</sup><br>% | Round 1<br>no. (%) | Round 2 no.<br>(%) | Round 3 no.<br>(%) |
| --- | --- | --- | --- | --- |
| Invited to round |  | 7166 | 6242 | 5506 |
| Participated in study round |  | 5501 (76.8) | 5603 (89.8) | 4806 (87.3) |
| Gender |  |  |  |  |
| Female |  | 3451 (62.7) | 3509 (62.6) | 3040 (63.3) |

|  | **PS Strata<sup>a</sup>**<br>**%** | **Round 1**<br>**no. (%)** | **Round 2 no.**<br>**(%)** | **Round 3 no.**<br>**(%)** |
| --- | --- | --- | --- | --- |
| Male |  | 1996 (36.3) | 2036 (36.3) | 1721 (35.8) |
| Other |  | 51 (0.9) | 52 (0.9) | 40 (0.8) |
| Missing |  | 3 (0.1) | 6 (0.1) | 5 (0.1) |
| Sex |  |  |  |  |
| Female | 51.8 | 3502 (63.7) | 3559 (63.5) | 3080 (64.1) |
| Male | 48.2 | 1996 (36.3) | 2042 (36.4) | 1725 (35.9) |
| Missing |  | 3 (0.1) | 2 (0) | 1 (0) |
| Age |  |  |  |  |
| 18-29 | 24.2 | 476 (8.7) | 378 (6.7) | 286 (6) |
| 30-44 | 29.0 | 1768 (32.1) | 1656 (29.6) | 1330 (27.7) |
| 45-64 | 30.0 | 2052 (37.3) | 2141 (38.2) | 1895 (39.4) |
| 65-74 | 10.2 | 928 (16.9) | 1077 (19.2) | 977 (20.3) |
| 75+ | 6.6 | 275 (5) | 345 (6.2) | 313 (6.5) |
| Missing |  | 2 (0) | 6 (0.1) | 5 (0.1) |
| Race/ethnicity |  |  |  |  |
| African American/Black | 17.4 | 268 (4.9) | 180 (3.2) | 146 (3) |
| American Indian/Alaska<br>Native/Other | 0.5 | 123 (2.2) | 114 (2) | 84 (1.7) |
| Asian/Pacific Islander | 20.3 | 862 (15.7) | 763 (13.6) | 668 (13.9) |
| Hispanic | 22.4 | 860 (15.6) | 637 (11.4) | 527 (11) |
| Two or more races | 3.8 | 500 (9.1) | 400 (7.1) | 334 (6.9) |
| White | 35.6 | 2886 (52.5) | 3503 (62.5) | 3041 (63.3) |
| Missing |  | 2 (0) | 6 (0.1) | 6 (0.1) |
| Educational attainment |  |  |  |  |
| College degree or more | 47.0 | 4912 (89.3) | 5101 (91) | 4404 (91.6) |
| Less than college degree | 53.0 | 552 (10) | 496 (8.9) | 399 (8.3) |
| Missing |  | 37 (0.7) | 6 (0.1) | 3 (0.1) |
| Current annual household income,<br>USD |  |  |  |  |
| <\$100K | 62.9 | 2081 (37.8) | 2075 (37) | 1779 (37) |
| ≥\$100K | 37.1 | 3192 (58) | 3347 (59.7) | 2866 (59.6) |
| Missing |  | 228 (4.1) | 181 (3.2) | 161 (3.3) |

Abbreviations: PS, Poststratification; USD, United States Dollar.
<sup>a</sup> Population percentages from synthetic poststratification tables generated from American Community Survey (ACS) and Public Use Microdata Sample data. Each cell contains the percentage of population age >18. Gender is not available in ACS data.

**Fig 1.**
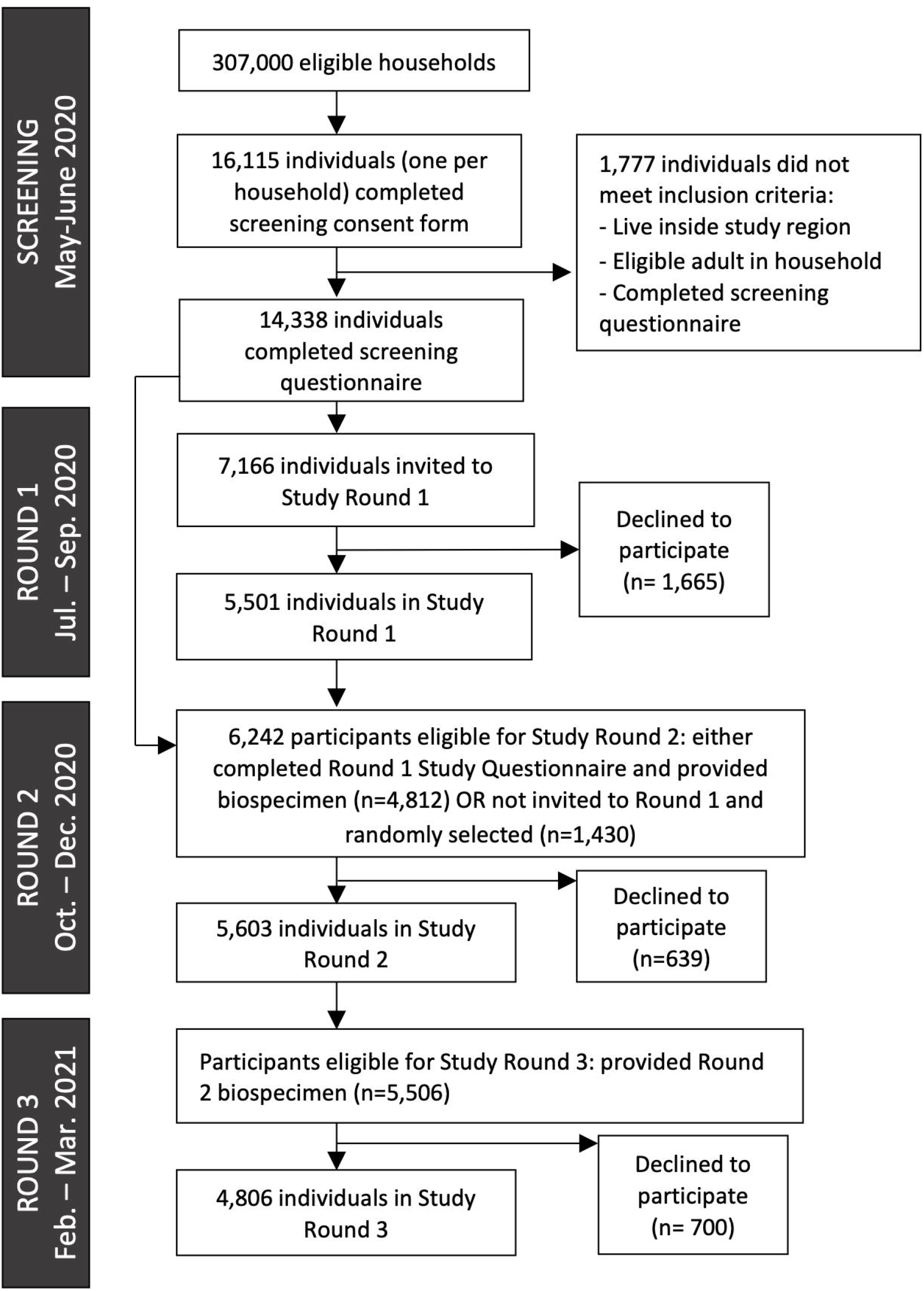
Study flow chart.

Of those who completed the questionnaire, 87.3%, 95.3%, and 96.6% provided DBS and 93.6%, 98.1% and 98.5% provided nasal swabs in rounds 1, 2, and 3, respectively (Table 2). Antibodies against the SARS-CoV-2 spike protein were detected in 29 (0.6%) and 33 (0.6%) of DBS in rounds 1 and 2, respectively. In Round 3, NC antibodies from natural infection alone were detected in 84 participants (1.8% of 4,806) and spike antibodies from natural infection or vaccination were detected in 1,452 participants (31.3% of 4,806). Viral infection was detected in less than three nasal swabs in each round. The proportion of participants reporting both being SARS-CoV-2 tested outside the study and testing positive increased over the study period: 10/1,030 (1.0%) participants reporting a positive COVID-19 result in Round 1, 19/2,059 (0.9%) in Round 2, and 53/1,892 (2.8%) in Round 3. Few participants met the criteria for being a COVID-19 probable case (<0.5%) (Table S-2).

**Table 2.** Prevalence of SARS-CoV-2 related outcomes among participants per study round.

|  | **Round 1** | **Round 2** | **Round 3** |
| --- | --- | --- | --- |
| Biospecimen collection period | Jul. 4, 2020 – Sep. 1, 2020 | Oct. 10, 2020 – Dec. 16, 2020 | Feb. 11, 2021 – Mar. 20, 2021 |
| N | 5501 | 5603 | 4806 |
| Provided DBS, no. (%) | 4801 (87.3) | 5340 (95.3) | 4641 (96.6) |
| Provided nasal swab, no. (%) | 5148 (93.6) | 5499 (98.1) | 4735 (98.5) |
| SARS-CoV-2 spike antibody status<sup>a</sup>, no. (%) |  |  |  |
| Non-reactive | 4641 (96.7) | 5275 (98.8) | 3157 (68.0) |
| QNS | 131 (2.7) | 31 (0.6) | 32 (0.7) |
| Reactive | 29 (0.6) | 33 (0.6) | 1452 (31.3) |
| SARS-CoV-2 nucleocapsid antibody status<sup>b</sup>, no. (%) |  |  |  |
| Non-reactive |  |  | 4181 (90.1) |
| QNS |  |  | 376 (8.1) |
| Reactive |  |  | 84 (1.8) |
| SARS-CoV-2 viral status, no. (%) |  |  |  |
| Invalid/Inconclusive | 4 (0.1) | 5 (0.1) | 7 (0.1) |
| Not Detected | 5143 (99.9) | 5491 (99.9) | 4725 (99.8) |
| Positive 2019-nCoV | 1 (0.0) | 3 (0.1) | 3 (0.1) |
| Self-reported positive COVID-19 test, no. (%) |  |  |  |
| No | 1020 (18.5) | 2059 (36.7) | 1892 (39.4) |
| Yes | 10 (0.2) | 19 (0.3) | 53 (1.1) |
| Not tested | 4471 (81.3) | 3525 (62.9) | 2861 (59.5) |
| Probable COVID-19 case<sup>c</sup>, no. (%) |  |  |  |
| No | 4800 (87.3) | 4938 (88.1) | 4419 (91.9) |
| Yes | 21 (0.4) | 10 (0.2) | 8 (0.2) |
| Not reported | 680 (12.4) | 655 (11.7) | 379 (7.9) |

Abbreviations: DBS, dried blood spot; QNS, quantity not sufficient.
<sup>a</sup> Antibody assays specific to spike antibodies detect presence of antibodies induced by SARS-CoV-2 natural infection or vaccination.
<sup>b</sup> Antibody assays specific to nucleocapsid antibodies detect presence of antibodies induced by SARS-CoV-2 natural infection.
<sup>c</sup> Defined by the Council for State and Territorial Epidemiologists.

### Population adjusted SARS-CoV-2 outcome prevalence in study region

Population adjusted SARS-CoV-2 seroprevalence, self-reported COVID-19 test positivity, and probable COVID-19 cases are reported in Table 3. Overall, populated-adjusted SARS-CoV-2 natural infection seroprevalence was low across the study region: Round 1 (July-September 2020) 1.03% (95% CI 0.50-1.96), Round 2 (October-December 2020 1.37% (0.75-2.39), and Round 3 (February-March 2021) 2.18% (95% CI 1.48-3.17). In Round 3 the populated-adjusted seroprevalence of COVID-19 vaccination was 21.64% (95% CI: 19.2, 24.34). Models incorporating sensitivity and specificity of the antibody assays yielded lower SARS-CoV-2 seroprevalence estimates in Round 1, similar estimates in Rounds 2 and 3, and a higher estimate of COVID-19 vaccine seroprevalence in Round 3. Population adjusted self-reported test positivity to SARS-CoV-2 was similar in Rounds 1 (1.11%, 95 CI: 0.39-2.40) and 2 (1.29%, 95% CI: 0.55, 2.17) to seroprevalence estimates and increased to 4.58% (95% CI: 2.56-7.64) in Round 3. Population-adjusted prevalence of being a COVID-19 probable case was <1% across rounds (Figure S-2).

**Table 3.**
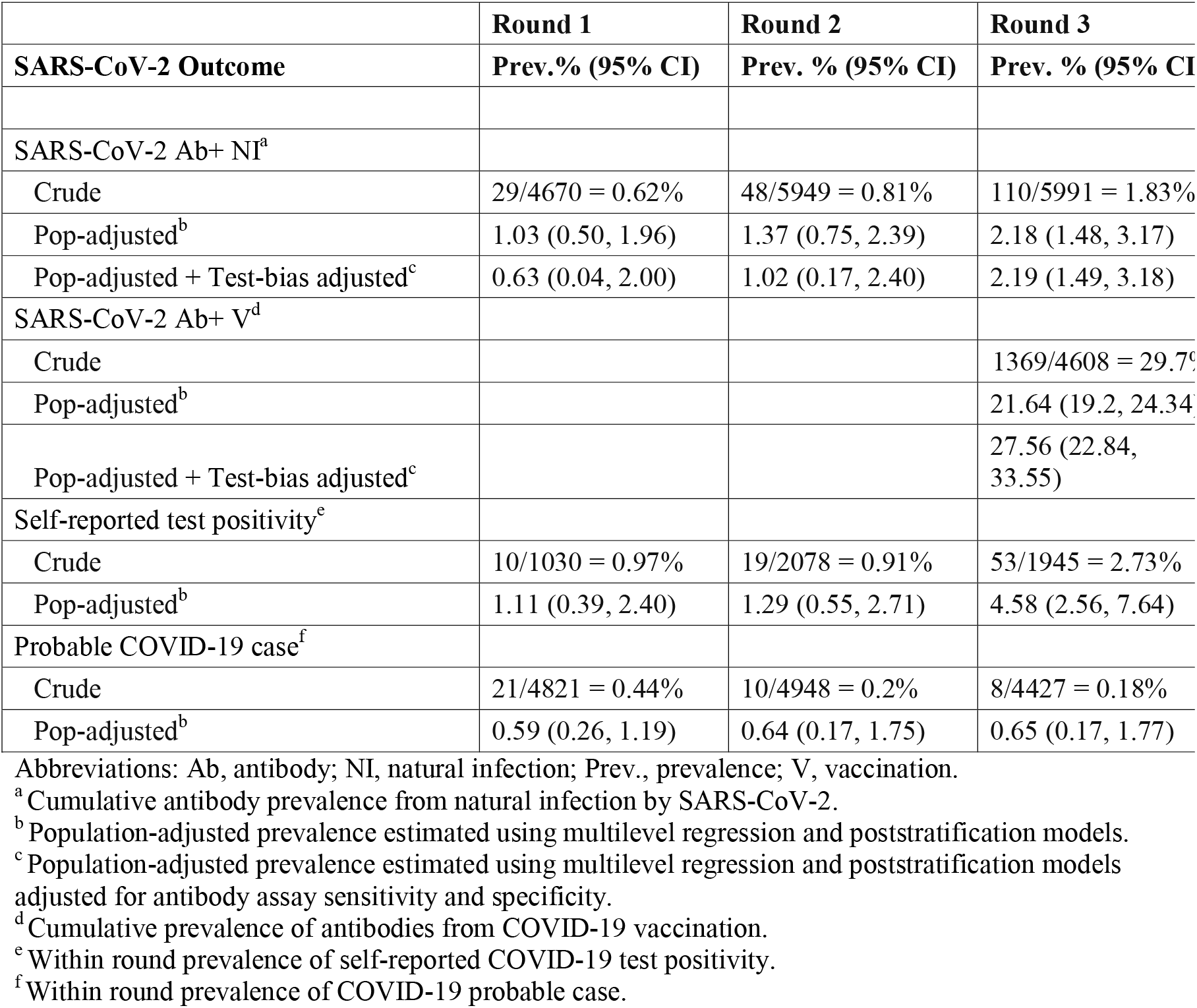
**Crude and population-adjusted prevalence (%) and 95% credible intervals of SARS-CoV-2 outcomes within the study region**.

### SARS-CoV-2 seroprevalence and infection vary by geographic area

There was evidence for spatial differences in both populated-adjusted seroprevalence and self-reported test positivity (Fig 2). The northern areas (Richmond, San Pablo, Pinole, and Hercules) and southern areas (East Oakland) of the study region had higher seroprevalence than Berkeley, El Cerrito, and North/Downtown Oakland. Self-reported test positivity was also higher in the northern and southern areas. These trends were consistent across study rounds.

**Fig 2.**
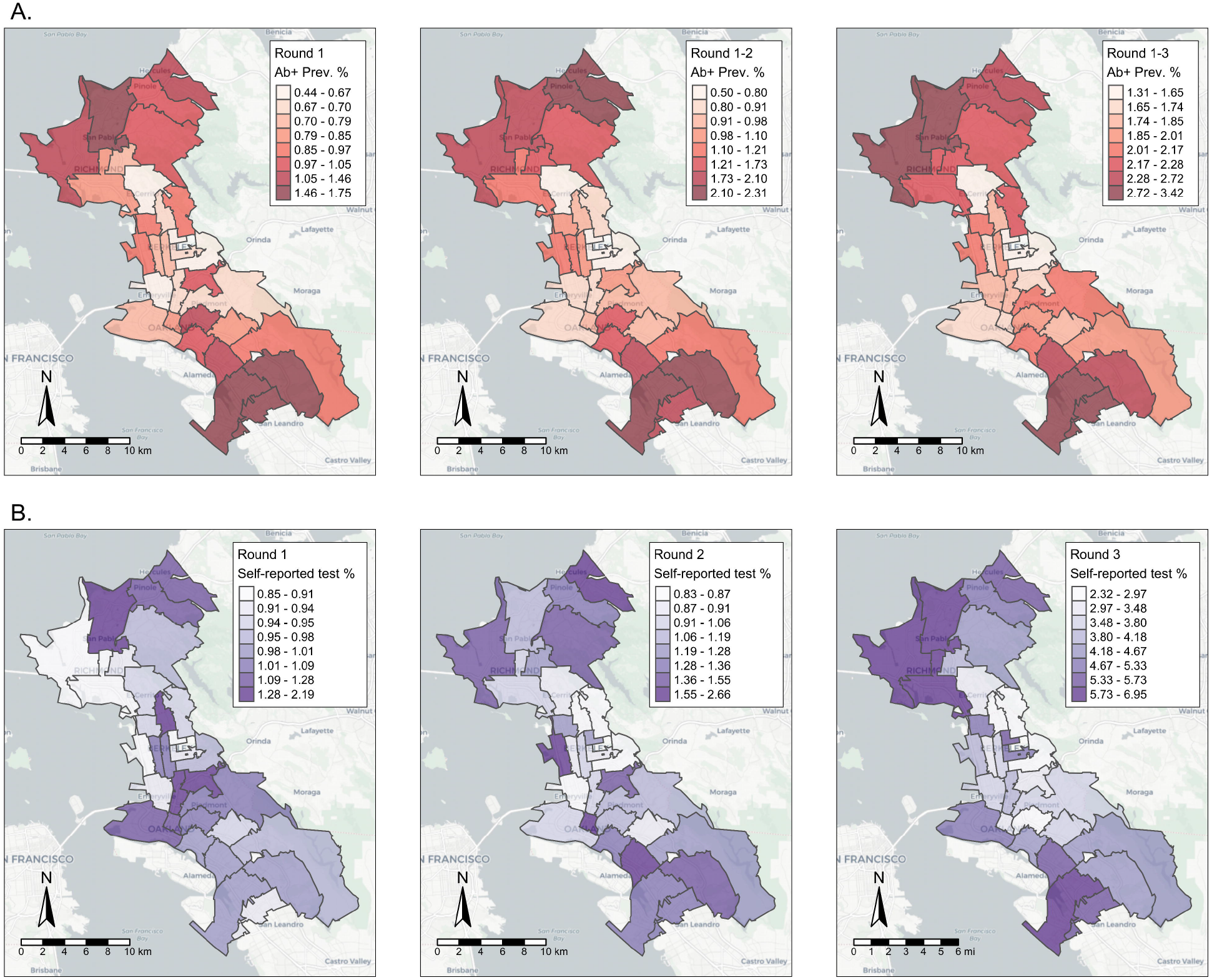
Population-adjusted prevalence of SARS-CoV-2 outcomes within study region ZIP codes. (A) Cumulative seroprevalence of SARS-CoV-2 antibodies to natural infection. (B) Prevalence of self-reported COVID-19 test positivity across the study region. Data collected in three rounds: Round 1, July-September 2020; Round 2, October-December 2020), and Round 3, February-March 2021.

### Population adjusted seroprevalence and self-reported test positivity prevalence within subgroups

Differences in populated-adjusted seroprevalence were observed among demographic groups (Fig 3). In Round 1, those aged less than 45 years had higher likelihood of antibody positivity than those aged 45 or older, but this relationship reversed over time. In general, non-Whites consistently had a higher likelihood of antibody positivity than Whites. The likelihood of antibody positivity also differed by household income and educational attainment, with those reporting a household income <$100,000 and less than a college degree having a higher likelihood of antibody positivity than their comparison groups. Those in households with more than four people had a higher likelihood of antibody positivity than those in households with four or less people. There was no clear relationship between sex and populated-adjusted seroprevalence. Similar relationships were seen between demographic groups and self-reported test prevalence (Figure S-3).

**Fig 3.**
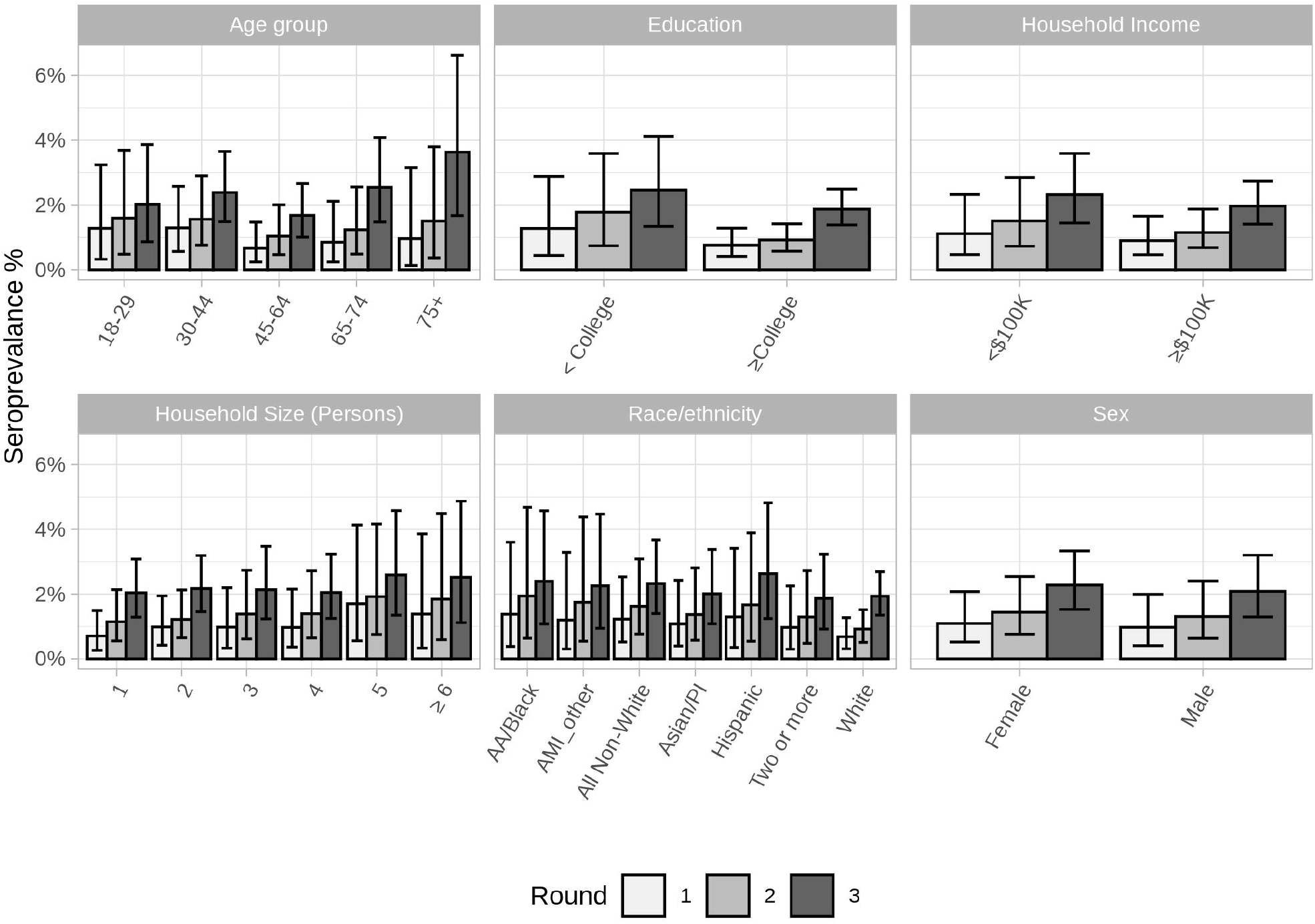
Cumulative seroprevalence of SARS-CoV-2 antibodies to natural infection among demographic groups. A) Sex, B) Age, C) Race/ethnicity, D) Income, E) Educational attainment, and F) Household size. Abbreviations: AA, African American; AMI, American Indian or Alaskan Native; PI, Pacific Islander.

### COVID-19 vaccination seroprevalence differs by race/ethnicity and age

In Round 3, populated-adjusted COVID-19 vaccination seroprevalence was lower in all non-White groups compared to Whites, (White vs. Non-White, PD: -4.35%, 95% CI: (−9.32, -0.35), Table 4, Figure S-4). The difference between racial/ethnic groups was largest among older participants; non-Whites had a lower COVID-19 vaccination seroprevalence compared to Whites among those aged between 65-74 (PD: -11.67%, 95% CI: (−20.98, -2.49)) and those aged 75 or older (PD: -10.15%, 95% CI: (−22.25, 0.43)).

**Table 4.** **Population-adjusted prevalence of antibodies from COVID-19 vaccination in Round 3 within race/ethnicity and age groups and prevalence differences between non-Whites and Whites**.

|  | Within group<br>Prev. % (95% CI)<sup>a</sup> | Non-Whites<br>compared to Whites<br>PD % (95% CI)<sup>b</sup> |
| --- | --- | --- |
| Race/ethnicity |  |  |
| White | 25.33 (23.14, 27.65) | Ref |
| African American/Black | 19.69 (13.47, 26.13) | -5.65 (-12.17, 0.76) |
| American Indian/Alaska<br>Native/Other | 18.87 (12.32, 25.95) | -6.46 (-13.3, 0.51) |
| Asian/Pacific Islander | 21.34 (17.24, 26.13) | -4 (-8.4, 0.93) |

|  | **Within group** | **Non-Whites**<br>**compared to Whites** |
| --- | --- | --- |
|  | **Prev. % (95% CI)<sup>a</sup>** | **PD % (95% CI)<sup>b</sup>** |
| Hispanic | 18.64 (13.74, 24.4) | -6.69 (-11.83, -0.81) |
| Two or more races | 15.74 (11.85, 19.94) | -9.6 (-13.88, -5.28) |
| All non-White | 19.6 (16.16, 23.33) | -4.35 (-8.32, -0.35) |
| Age group, years |  |  |
| 18-29 | 15.04 (10.92, 19.81) | -3.2 (-9.14, 2.23) |
| 30-44 | 16.53 (13.84, 19.57) | -1.94 (-6, 2.74) |
| 45-64 | 16.6 (13.99, 19.61) | -1.04 (-5.44, 3.89) |
| 65-74 | 45.74 (40.72, 50.75) | -11.67 (-20.98, -2.49) |
| 75 + | 54.01 (46.12, 61.79) | -10.15 (-22.25, 0.43) |

Abbreviations: CI, credible interval; PD, prevalence difference; Prev, prevalence.
<sup>a</sup> Population adjusted seroprevalence in round 3 estimated using multilevel regression and poststratification (MRP); Seroprevalence and 95% CI are the mean and 2.5% and 97.5% quantiles of a posterior distribution respectively. Assay used detects antibodies from natural SARS-CoV-2 infection or from COVID-19 vaccination.
<sup>b</sup> Population adjusted COVID-19 seroprevalence difference in round 3 between each race/ethnicity group and Whites among race/ethnicity and between non-Whites and Whites within each age group. PDs estimated using MRP, and the PDs and 95% CIs are the mean and 2.5% and 97.5% quantiles of a posterior distribution, respectively.

### Mask-wearing and association between high-risk vs. low-risk behavior and seroprevalence

More than 99% of participants reported ever wearing a mask 99% reported wearing a mask during leisure and exercise activities, >91% reported wearing a mask at work, and >88% reported wearing a mask while shopping (Table S-3). After clustering participants into “low-risk” and “high-risk” groups according to self-reported mitigation behaviors, most participants were considered low-risk across the study rounds (70%, 82%, and 77%, respectively; Figure 5A). Behaviors with the largest differences in high-versus low-risk behavior were reporting “yes” to: left home for work, medical/healthcare, care of relative, or other; worked with potential COVID-19 contact; attended gathering; and traveled to county outside of residence within last two weeks (Figure 5B). Age, race/ethnicity, and education were all associated (*P*<0.001) with high-vs. low-risk groups (Table S-4), but we did not observe strong evidence for an association between mitigation behavior and either seroprevalence or self-reported test positivity (Table 5). Although point estimates indicate that “low-risk” behavior was associated with lower seroprevalence.

**Table 5.** **Association of high-risk vs. low-risk mitigation behavior with seroprevalence and self-reported test positivity and within each study round**.

|  | MOA<sup>a</sup> | Round 1 | Round 2 | Round 3 |
| --- | --- | --- | --- | --- |
| SARS-CoV-2 antibody prevalence<sup>b</sup> | PD % (95% CI) | -0.21 (-1.4, 0.79) | 0.02 (-1.59, 1.03) | -0.46 (-2.23, 1.00) |
|  | PR (95% CI) | 0.93 (0.34, 2.03) | 1.4 (0.32, 3.93) | 0.88 (0.45, 1.57) |
| Self-reported test positivity | PD % (95% CI) | 0.53 (-0.73, 2.24) | 0.11 (-2.12, 1.6) | -1.11 (-5.26, 2.18) |
|  | PR (95% CI) | 2.17 (0.54, 6.72) | 1.56 (0.33, 4.65) | 0.87 (0.41, 1.64) |

Abbreviations: CI, credible interval; MOA, measure of association; PD, prevalence difference; PR, prevalence ratio
<sup>a</sup> Populated adjusted prevalence estimated using multilevel regression and poststratification models; point estimate is the mean of a posterior distribution of the parameter, 95% credible interval estimated from 2.5% and 97.5% quantiles of a posterior distribution.
<sup>b</sup> Defined as having detectable SARS-CoV-2 antibodies within a study round.

## Discussion

In the current study, we investigated individual-level characteristics and behaviors that contributed to SARS-CoV-2 related outcomes, including seroprevalence and self-reported infection, in a large, population-based sample of over 5,500 participants from 12 East Bay (Northern California) cities. During three time periods from July 2020 to March 2021, we estimated the population-adjusted prevalence of SARS-CoV-2 outcomes across the study region and within strata of age, sex, race/ethnicity, ZIP code, and household size. We then characterized behaviors to mitigate transmission of SARS-CoV-2 and their association with related outcomes. Overall, prevalence of SARS-CoV-2 outcomes for natural infection were low which may be attributable to the high percentage of mask-wearing and other risk-mitigating behaviors among our participants. COVID-19 vaccination seroprevalence estimates in Round 3 was greater than 20%, with non-Whites having much lower seroprevalence than Whites.

Despite the low overall SARS-CoV-2 seroprevalence and infection observed in our study, ZIP code of residence, age, racial/ethnic identification, education, and income all contributed to differences in seroprevalence. Specifically, non-Whites, those without a college degree, and lower income households, had higher seroprevalence. Further, ZIP codes with higher proportions of Spanish speakers had higher populated-adjusted seroprevalence estimates (Figure S-5). These differences persisted despite the high rates of mask wearing reported by our study sample, further adding to strong evidence that the risk of COVID-19 is distributed unequally and that structural inequities play an important role in COVID-19 risk.[20–23] Moreover, during Round 3 (February-March 2021), COVID-19 vaccines were widely available across the study region. We found that Whites had much higher prevalence of antibodies from COVID-19 vaccination in Round 3, than non-Whites, as reported elsewhere.[3,24] Notably, this difference was largest among those aged 65 or older. Vaccine hesitancy does not explain these findings. In Round 3, <8% of participants reporting not receiving a COVID-19 vaccine within each racial/ethnic group were hesitant to vaccination and of the 81 participants reporting hesitancy, 63 (77.8%) were unsure of their vaccination plans. These findings demonstrate that in the first few months of vaccine availability in the Bay Area, large disparities in vaccination rates by race/ethnicity were observed among older persons. Furthermore, Whites, the group with the lowest prevalence of SARS-CoV-2 infection, were more likely to be vaccinated, underscoring the inequities that exist surrounding the coronavirus pandemic.

Another key finding was that almost all participants reported wearing masks. This contrasts with models of mask usage reported by the Institute for Health Metrics and Evaluation which reported mask usage ranging between 75-82% from December 2020 through March 2021 in California.[25,26] Mask wearing is one the most effective behaviors for controlling community spread of SARS-CoV-2 infection.[27] The high rate of mask usage by study participants may partially explain why we did not detect a differences between high-risk and low-risk mitigation behavior and SARS-CoV-2 prevalence, and partially explain why our estimates of SARS-CoV-2 seroprevalence and self-report test positivity were lower than public case reports. However, a recent study estimating SARS-CoV-2 seroprevalence from blood donors reported seroprevalence in the Bay Area region of California that were similar to our estimates through December 2020.[28]

A major strength of this study was the longitudinal design and collection of individual-level data, including biospecimens for antibody and virus testing, which is challenging but much needed in current studies of the pandemic. Comprehensive data on social distancing, self-quarantine, mask wearing, working from home, and other transmission mitigation efforts are also needed to inform current and future prevention strategies. At-home collection of biospecimens, including DBS for antibody testing, made regular testing without in-person interaction possible. This was a critical feature, particularly early in the pandemic, when recommendations were to travel only for essential purposes and to limit in-person interactions. At-home sample collection was used to obtain more than 30,000 biospecimens and is a feasible approach for large populations and geographic regions.

One limitation of this study was the under-representation of certain demographics in our sample. Supplementary mailings of recruitment postcards in Spanish were sent to residences in ZIP codes with high proportions of Spanish speaking households. We also placed recruitment flyers in local grocery stores and conducted outreach to community organizations, local government officials, and school districts in the study region. Despite these efforts, non-Whites, males, lower income households, and individuals with lower education and from lower socioeconomic ZIP codes were underrepresented in our sample. Unhoused individuals were also not captured in our sample. This was important given evidence that individuals who identify as Hispanic or Black, and other underrepresented groups, are at the highest risk for COVID-19.[23] These discrepancies may have led to underestimation of SARS-CoV-2 seroprevalence, although the MRP models may have partially minimized this by pooling information across similar observed characteristics in the sample data. Our seroprevalence estimates were consistent with COVID-19 case prevalence reported by public health agencies in ZIP codes with high response rates (Figure S-6).[29] Additionally, there may be unmeasured confounding from variables not included in the analyses and self-selection bias from some participants in COVID-19 research who may be more fervent adherers to COVID-19 public health measures. Finally, some individuals do not seroconvert after infection or vaccination.

Our results underscore the substantial and persistent inequities that exist surrounding the coronavirus pandemic. Non-Whites, lower-income, and lower-educated individuals had the highest SARS-CoV-2 seroprevalence. This disparity in seroprevalence was observed despite near universal rates of mask wearing in our sample. We also observed large differences in COVID-19 vaccination seroprevalence between racial and ethnic groups. More work must be done to address these disparities and inequities, such as allocation of resources for high-risk communities and strategies to mitigate the structural barriers posed by social and structural determinants of health.

## Data Availability

De-identified individual participant data that underlie the results reported in this article (text, tables, figures may be shared for up to 36 months following publication after investigators whose proposed use of the data has been approved by an independent review committee. For individual participant data analysis or meta-analysis, proposals should be directed to Lisa Barcellos and Lynn Hollyer. Requests will be reviewed by an independent review committee and the UC Berkeley Institutional Review Board. Data may be shared upon approval. Data requestors will need to sign a data access agreement.

## Acknowledgments

We want to acknowledge the contributions of the following individuals to this study: Ella Parsons, Jordan Keen, Janine Solomon, Jose Salinas, Kevin Duong, Joseph Egbunikeokye, Maya Talavera, Riya Shrestha, Colin Warnes, José Victor Zambrana, Nicholas Lo, Parnal Narvekar, Fausto Bustos, Gregorio Dias, Reinaldo Mercado-Hernandez, Julia Huffaker, Raymond Montes, Alexandra Zermeno, Alejandra Zeiger, William Dow, Michael Lu, Lila Krop, Kelly Lam, Yan Zhang, Sarah Folkmanis, Sophie Zhai, Dingjun Chen, Ruben Vargas Ethan Garcia, Oliver Li, Manisha Sahoo, Raina Walencewicz, Sophia Wang, Antonia Gibbs, Amrita Ramanathan, Catherine Livelo, Taylor Worley, Amanda Tanaka, Savinnie Ho, Jane Liu Ryan Allen, Sofia Soltero, Victoria Van Metter, Madeleine Fraix, Allie Coyne, Subeksha Sharma Lydia Yu, Shreeya Garg, Sanjeet Paluru, Malika Saxena, Talia Panadero, Ayra Rahman, Joshua Calangian, Dharaa Upadhyaya, Sophia Kemp, Ruhi Parikh, Amy Rich, Sophie Manoukian, Nola Vu, Crystal Nguyen, Jordyn Pinochi, Alma Kuc, Siri Ylenduri, Manvir Kaur, Angikaar Chana, and Sannidhi Sarvadhavabhatla, Benjamin T. Auch, Dinesha Walek, Evan Forsberg, Jerry Daniel, Veronica Tonnell, Ji Hyun (Jay) Kim, Mary Nieuwenhuis. Creative Testing Solutions: Valerie Green, Sherri Cyrus, Phillip Willamson, Brett Hirsch, Paul Contestable, Mars Stone, Joe Derisi, Emily Crawford, Emily Ahlvin, Armando Diaz, and Favianna Rodriquez.

## Funding

This study was supported by Open Philanthropy Projects, Fast Track grants from Emergent Ventures, the UC Berkeley Innovative Genomics Institute, and the UC Berkeley School of Public Health Center for Population Health.

## Conflict of interest

JAL reports receipt of grants unrelated to this study from Pfizer and Merck, Sharpe & Dohme, and consulting fees from Pfizer, Merck, Sharpe & Dohme, VaxCyte, and Kaiser Permanente. All other authors report no disclosures.

## Author Contributions

LB, EH, JC, MB, JAL, KB, CA, and MH conceived of the study. IF, XS, CA, and MH were responsible for data curation. Formal data analysis was performed by CA, OS, and SF. Financial support for the work presented in this study was acquired by LB and EH. Data acquisition, including designing of surveys, design and construction of biospecimen collection kits, FedEx mailings, receiving of biospecimens mailings from participants, and communication with participants was performed by LL, IF, MH, HQ, DQ, AB, JS, XS, LB and MH. Viral and antibody testing was performed and/or supervised by KB, BA, MW, DM, MM, SM, YS, HQ, and DQ. Sampling design was conceived of by JAL and CA, and models for prevalence analyses were developed by CA and SW. Management and coordination for EBCOVID study was primarily the responsibility of LF and EH, with support from JC and MH. LF and EH research facilities were the primary source of study and laboratory materials. Computing resources were provided by LF. Implementation of NIMBLE models was performed by CA with support from SW and SF. IF was responsible for management and implementation of REDCap surveys and overall system administration of computing resources necessary for the research. LF and EH supervised all research performed as part of the EBCOVID study. LB, EH, CA, MH, and IF had full access to and verified the study data. CA, OS, and MH created all figures in the manuscript. CA and MH wrote and prepared the initial draft of this manuscript. All authors contributed to interpreting the results of analyses and critically editing and revising the manuscript.

## Notes

### Competing Interest Statement

Joe Lewnard reports receipt of grants unrelated to this study from Pfizer and Merck, Sharpe & Dohme, and consulting fees from Pfizer, Merck, Sharpe & Dohme, VaxCyte, and Kaiser Permanente. All other authors declare no competing interests.

### Funding Statement

This study was funded by grants from Open Philanthropy, Fast Grants, Mercatus Center, the UC Berkeley Innovative Genomics Institute, and the UC Berkeley Center for Population Health.

### Author Declarations

The study was approved by the University of California, Berkeley Committee on Protection of Human Subjects (Protocol #2020-03-13121).

### Summary of Updates

Reduced number of figures in manuscript and moved to supplement. Minor edits to text.

