## Supplementary Methods, Tables, and Figures for "Health inequities in SARS-CoV-2 infection, seroprevalence, and COVID-19 vaccination: Results from the East Bay COVID-19 study"

### 1 SARS-CoV-2 viral detection

Nucleic acid extraction to obtain (potential) viral RNA from study participants was performed in collaboration with the University of Minnesota Genomics Center. Briefly, nasal/nares swab samples provided by study participants in collection tubes with a stabilizing solution were first heat-inactivated and transferred to a 96-well plate for further processing to obtain RNA. Solid Phase Reversible Immobilization (SPRI) was used for nucleic acid extraction as previously described.[1] Briefly, Anterior nares (AN) swab samples in 3 mL DNA/RNA Shield (Zymo) were first heat-inactivated at 56°C for 20 minutes. 200 µL of the resulting solution was transferred to a 96-well deep well plate containing 150 uL of Lysis Buffer (4.67M GuHCl, 5.8 mM TCEP, 23.3 mM EDTA, 23.3 mM Tris, pH 7.0, 0.23 % (v/v) Igepal CA-630, 100 units/mL Proteinase K) and pipette-mixed 10 times. The plate was sealed and again incubated at 56°C for 20 minutes.

Following lysis/digestion, 350 µL of a SPRI bead solution (1:50 dilution of 3X washed Cytiva Sera-Mag SpeedBeads™ 65152105050250, 18% (w/v) PEG-8000, 1M NaCl, 10 mM Tris-HCl, 1 mM EDTA) was added to the lysate and pipette-mixed 10 times, incubated at room temperature for 10 minutes, and pipette-mixed an additional 10 times. Following incubation and mixing, the plate was placed on a magnet and the magnetic beads were allowed to pellet. After, the supernatant was removed, and the beads were washed twice with 800 µL freshly prepared 80% ethanol. Beads were allowed to dry off-magnet for 10 minutes at room temperature, and then were resuspended with 50 µL nuclease-free H20. After magnetizing again, the clear supernatant was removed to a fresh plate, and this solution was used as input for RT-qPCR.

This RNA was used as input for reverse transcription quantitative real-time polymerase chain reaction (RT-qPCR). Three separate RT-qPCR reactions were set up for each study participant sample in a 384-well plate, and PCR was performed using CDC-specified primers and Taqman probes for viral N1 and N2 genes, and human RNase P (https://www.cdc.gov/coronavirus/2019-ncov/downloads/rt-pcr-panel-primer-probes.pdf) as described.[2] Following PCR thermocycling steps, the amplification results for each run were manually inspected, and results were interpreted to determine whether there was evidence of coronavirus infection for each participant (amplification cycle threshold for N1 or N2 gene $\leq 40$).

Lysis Buffer

- 4.67M GuHCl
- 5.8 mM TCEP
- 23.3 mM EDTA
- 23.3 mM Tris, pH 7.0
- 0.23 % (v/v) Igepal CA-630
- 100 units/mL Proteinase K

Beads

- 1:50 dilution of 3X washed Cytiva Sera-Mag SpeedBeads™
- 18% (w/v) PEG-8000

- 1M NaCl
- 10 mM Tris-HCl
- 1 mM EDTA

### 2 SARS-CoV-2 antibody detection

Antibody testing was performed using dried blood spots (DBS) provided by study participants.

#### 2.1 Quality Control of Dried Blood Spots

Participant's DBS sample cards (containing six discs) were assessed for quality. We evaluated each DBS disc on a scale from 0-3, with 0 representing a blank DBS, 1 representing an incompletely filled DBS on one side, 2 representing a DBS card saturated fully on one side, and 3 representing a DBS fully saturated on both sides. Participants that provided at least two DBS discs with a cumulative score of two or higher were processed in our serological assays. Samples that did not meet this criterion were labelled as "Quantity Not Sufficient" (QNS).

#### 2.2 Reconstitution of Blood Spots

Two DBSs were cut from disks and placed in 2mL screwcap tubes. 500uL of elution buffer was added into tube and vortexed for 20 seconds. If spots were limited, one spot was eluted in 250uL of elution buffer instead. DBS was incubated in elution buffer overnight at 4°C, then spun down at 10,600 g for 10 min at 4°C. DBS eluate was then transferred to a fresh tube and stored at 4°C until analysis.

#### 2.3 Antibody testing

For each study round, two high quality DBS discs from each participant were rehydrated by soaking them in a reconstitution solution and were processed for testing. Over the course of the study, three assays were used: Ortho VITROS® Anti-SARS-CoV-2 Total Ig test (ORTHO), anti-Spike IgG ELISA Assay (ELISA), and Roche NC Total Ig (ROCHE).

##### *2.3.1 Ortho VITROS® Anti-SARS-CoV-2 Total Ig test*

DBS eluates were tested on the Ortho VITROS® Anti-SARS-CoV-2 Total test according to the manufacturer's instructions described briefly. The Ortho VITROS Anti-SARS-CoV-2 total (CoV2T, Ortho-Clinical Diagnostics, Inc) was used to detect total (IgG, IgM, and IgA) antibodies to SARS-CoV-2 Spike S1 protein. Briefly, DBS eluates or plasma samples were loaded on Ortho VITROS XT-7200 or 3600 instruments (Ortho-Clinical Diagnostics, Inc) and programmed for the CoV2T test following the manufacturer's instructions. The S1 antigens coated on the assay wells bind S1 antibodies from human serum which, in turn, bind to a secondary HRP-labeled S1 antigen in the conjugate reagent forming a sandwich. The addition of signal reagent containing luminol generates a chemiluminescence reaction that is measured by the system and quantified as the ratio of the signal relative to the cut-off value generated during calibration. A S/CO ≥1 was considered positive.

##### *2.3.2 anti-Spike IgG ELISA Assay*

DBS eluates and plasma were evaluated for the presence of IgG against SARS-CoV-2 Spike (S) and using an in-house direct ELISA as previously described.[3] Briefly, SARS-CoV-2 antigens were coated in 96-well Nunc Maxisorp ELISA plates (ThermoFisher) overnight at 4°C. Plates were then blocked in 2.5% non-fat dry milk in PBS for 2h at 37°C. Plates were then washed 3x with PBS and 100uL of DBS eluate in 1% non-fat dry milk and incubated at 37°C for 1h. Plates were then washed 5 times with 0.05% PBS-Tween-20 and 100uL of goat-α-human IgG HRP secondary antibody (Fisher) diluted in 1% non-fat dry milk was added. After incubating for 1h at 37°C, plates were washed 5 times in 0.05% PBS-Tween-20 and once in PBS. Plate was developed with TMB (3,3',5,5'-Tetramethylbenzidine) for exactly 5 minutes and reaction was stopped with 2M $H_2SO_4$. Plates were read on a plate reader at 490nm.

##### *2.3.3 Roche NC Total Ig*

The Roche Elecsys Anti-SARS-CoV-2 immunoassay (Roche NC) was run on the Cobas e441 analyzer (Roche Diagnostics) to detect antibodies against the SARS-CoV-2 nucleocapsid protein. DBS eluates were initially incubated with biotinylated and ruthenium-labeled SARS-CoV-2 recombinant nucleocapsid antigens and any antibody present in the solution is sandwiched between the two. Subsequently, streptavidin-coated microparticles are added to the mixture to bind the biotin. The magnetic particles drive the complexes to the electrode, where a chemiluminescent signal is emitted and measured as the ratio between the signal and the cut-off obtained during calibration. A S/CO $\geq$1 in plasma was considered positive while a S/CO $\geq$ 0.045 in DBS was considered positive.

#### 3 Recruitment and participants

Recruitment and selection of study participants was completed using a screening phase followed by a longitudinal study phase with three timepoints or "rounds". In the screening phase, all residential addresses within the East Bay cities and communities of Albany, Berkeley, El Cerrito, El Sobrante, Emeryville, Hercules, Kensington, Oakland, Piedmont, Pinole, Richmond, and San Pablo (n~307,000 residential households) were mailed an invitation to participate. The household member aged 18 or older with the next birthday was invited to complete a consent form and screening questionnaire. Spanish versions of study invitations and all study materials were also utilized.

Significant effort was spent recruiting individuals from ZIP codes with high percentages of Latino and non-white individuals. This included sending recruitment postcards in English and Spanish to households in ZIP codes with large percentages of Spanish speakers; outreach to local high schools to notify families of the recruitment effort; fliers in English, Spanish, Chinese, Korean, Vietnamese, and Tagalog sent to local city government officials and community benefit organizations; and outreach to local Spanish radio stations and markets.

Of the 16,115 residents who consented and completed the screening procedures between May-July 2020, 1,777 individuals did not meet the inclusion criteria and were excluded. Eligible participants were required to be the household member with the next birthday, live within the study region, and be willing to provide biospecimens and questionnaire responses, read and speak English or Spanish, have a valid email address, and have internet access. The target sample size for the study phase was 5,500 participants. To obtain a sample that resembled the racial and ethnic proportions reported in the 2018 American Community Survey (ACS) for the study region, we ranked screening participants for study inclusion. Black and/or Hispanic individuals were ranked the highest (n=1,556) followed by other non-white individuals (n=1,939). Order of inclusion for white individuals was randomly sampled. Individuals ranked between 1 and 5,500 were offered study enrollment, and non-respondents were replaced with next highest ranked individuals who had not yet been offered study entry.

Biospecimens and questionnaire data were collected during three rounds. Approximate dates for each round were July-September 2020, October-December 2020, and February-April 2021. For each round of data collection, individuals who had participated in the previous round were contacted to confirm their participation in the next round. If participation was declined, individuals from the pool of screening participants who had not yet participated in a study round were invited as needed. This resulted in 5,501 participants in Round 1, 5,603 participants in Round 2, and 4,806 participants in Round 3. This corresponded to participation rates of 76.8%, 89.8%, and 87.3% across the study round, respectively.

All participants provided their informed consent for the screening phase. All those participating in the study phase provided their informed consent for each study round. The study was approved by the University of California, Berkeley Committee on Protection of Human Subjects (2020-03-13121).

### 4 Data

#### 4.1 Study questionnaire

Participants were asked to complete an online questionnaire at the time biospecimens were collected for each round. The questionnaire included demographics, tobacco and alcohol use, income, employment, physical and mental health, medical conditions, healthcare coverage, COVID-19 symptoms, and past COVID-19 testing. Questions about the participant's household such as number of members were also collected. Screening and study questionnaire data were collected using REDCap.

#### 4.2 SARS-CoV-2 outcomes

For each study round, we considered three separate outcomes for analyses, (1) cumulative SARS-CoV-2 antibody positivity, (2) self-reported SARS-CoV-2 viral positivity from RT-qPCR testing, (3) and the standardized surveillance case definition of "probable COVID-19" based on self-reported symptoms, close contact with others who were infected, and viral testing. See below supplement methods for description of outcome definitions.

##### *4.2.1 SARS-CoV-2 seropositivity*

Broadly, SARS-CoV-2 seropositivity is defined as the presence of antibodies against SARS-CoV-2. Antibody assay testing algorithms implemented in this research were updated during the study to reflect changing conditions of the pandemic (Figure S-1, Table S-5). Briefly, in Round 1, samples with a signal/cutoff (S/C) value ≥1 on the Ortho-VITROS assay were considered to reflect previous infection by SARS-CoV-2. In Round 2, samples with both Ortho-VITROS S/C≥0.7 and ELISA optical density (OD) ≥0.34 were considered have antibodies from SARS-CoV-2 infection. During Round 3 vaccinations were widely available, therefore we targeted antibodies to the SARS-CoV-2 nucleocapsid (NC) protein which is specific to natural infection not vaccination: samples with Ortho-VITROS S/C≥0.7 and ROCHE S/C ≥ 0.0465 were considered to have antibodies from SARS-CoV-2 infection. Samples with Ortho-VITROS S/C≥0.7, ELISA optical density (OD) ≥0.34, and ROCHE S/C < 0.0465 were considered to have antibodies from COVID-19 vaccination alone.

To identify the cumulative seroprevalence of SARS-CoV-2 within our study region, cumulative seropositivity was defined as having detectable SARS-CoV-2 antibodies in the current and/or previous round(s) and cumulative seronegative was defined as not having detectable SARS-CoV-2 antibodies in the current and previous study rounds.

##### *4.2.2 Self-reported COVID-19 test positivity*

During each study round, participants were asked whether they had been tested for coronavirus (COVID-19) by a physician or medical professional since the previous data collection period and whether any test had been positive for SARS-CoV-2. Within each round, all participants who reported being tested were included the denominator; those who reported having a positive result for that test(s) were included in the numerator. Within a study round, self-reported COVID-19 test prevalence was defined as the proportion of those reporting a positive test out of all participants who reported having a test.

##### *4.2.3 Probable COVID-19*

We used the Council for State and Territorial Epidemiologists (CSTE) standardized case definition of "probable COVID-19" which is intended for public health surveillance purposes.[4] Individuals were classified as a "probable COVID-19" case if, within the previous 14 days, they reported being within six feet of a confirmed case of COVID-19 and reported at least two of the following symptoms: fever (measured or subjective), chills, rigors, myalgia, headache, sore throat, nausea, vomiting, diarrhea, fatigue, congestion, or runny nose. Within a study round, probable COVID-19 case prevalence was defined as the proportion of those being identified as a probable COVID-19 case out of all participants who provided responses to questions about recent close contact history and symptoms.

#### 5 Behaviors related to virus containment and mitigation

At each study round, participants were asked about physical distancing practices, recent close contacts with others, mask wearing, and behaviors and activities that might affect risk of SARS-CoV-2 or related outcomes. (See below for questions). To obtain a single binary measure of "mitigation behavior", we performed a latent class analysis (LCA) to classify participants into "high-risk" and "low-risk" behaviors classes, using the *poLCA* R package.[5–7] Sixty questions related to COVID-risk behaviors were grouped into 6 categories: 1) mask wearing, 2) leaving your home, 3) mode of transportation, 4) travel, 5) attendance at gatherings, and 6) occupational-related exposures (Table S-7). To create interpretable LCA classes (i.e., "high-risk" vs. "low-risk"), we first identified a subset of questions with each category that were the most representative of participant responses using similarity scores. For example, any masking wearing any of the queried situations (while at work, while traveling, while shopping, or while doing leisure activities outside) was representative of participants' mask wearing habits overall. A total of 15 variables across the six categories were identified using similarity scores and used for LCA classification. For the leaving home category, the most informative variables were leaving your home at all; leaving for either shopping, leisure, or exercise; and leaving for work, relative care, medical care, or another reason. Whether a participant used public or personal transportation was the most important feature of the transportation variables. Whether a participant traveled to countries outside the United States, states outside California, or counties outside of primary residence either since December 2019 or in the two weeks prior to completing the questionnaire were used from the travel questions. Whether a participant attended any gathering or attended a gathering with more than 10 people were both retained for the gathering group. Finally, whether a participant or a member of their household had potential contact with persons infected with COVID-19 at work informed work exposures.

### 6 Population-adjusted seroprevalence and other SARS-CoV-2 outcomes

#### 6.1 Summary of MRP

Bayesian multilevel regression and poststratification (MRP) was used to estimate population adjusted cumulative seroprevalence, self-reported SARS-CoV-2 viral positivity at each study round, and “probable COVID-19” at each study round. MRP is a regression-based method for estimating average and sub-average effects from survey data. MRP has been shown to perform better than survey weighting methods particularly when working with small sample sizes and sparse data. The hierarchical regression model achieves this by pooling information across similar observed characteristics in the model. Details of MRP are described here[8,9]. Briefly, the steps for MRP used in this study were as follows:

1. Identify demographic and geographic characteristics of interest in the study population and construct a table with the population size, $N_j$, for each stratum $J$ for all combinations of characteristics of interest.
2. Model outcome of interest as a function of demographic and geographic characteristics with the survey data using a multilevel regression model.
3. Predict probability of the outcome, $\hat{\pi}_j$, within each stratum $j$ using the fitted multi-level model.
4. Use $\hat{\pi}_j$ as post-stratification weights and aggregate predictions from strata to population or sub-population of interest.

$$\hat{\pi}_{pop} = \frac{\sum_{j \in J} N_j \, \hat{\pi}_j}{\sum_{j \in J} N_j}$$

6.2 Synthetic population and poststratification

In this study, demographic variables of interest were sex, age, race, Hispanic ethnicity, income, education, household size, and ZIP code. However, cross-tabulations for these variables were not available from a single data source for our study region. We used a method described by Leeman et al to generate a synthetic population using all of the variables described above for the post stratification step.[9] The American Community Survey (ACS) provides cross tabulations for sex, age, and race within ZIP Code Tabulation Areas (ZCTAs). Marginal distributions for Hispanic ethnicity, education, income, and household size were available at the ZCTA level. Synthetic populations were generated by combining sex, age, race, and ZCTA joint distributions and marginal distributions of Hispanic ethnicity, education, and income within ZCTA. Joint distributions at the individual level for these variables were extracted from the Public Use Microdata Sample (PUMS) for Public Use Microdata Areas within our study region. The joint distribution of these seven variables yielded the following number of strata,

$$J = J_{ZIP} \times J_{sex} \times J_{age} \times J_{race/eth} \times J_{edu} \times J_{income} \times J_{HHsize} 31 \times 2 \times 5 \times 6 \times 2 \times 2 \times 6 = 44{,}640 \;\; strata$$

6.3 Multilevel Regression model

At each study round, SARS-CoV-2 outcomes were modeled as a function of geographic and demographic characteristics using logistic regression,

$$y_i = logit^{-1}(\beta_0 + \beta_1 male_i + \beta_2 X_{zip[i]} + \alpha_{eth[i]} + \alpha_{age[i]} + \alpha_{edu[i]} + \alpha_{income[i]} + \alpha_{hh[i]} + \phi_{zip[i]} + \alpha_{zip.eth[i]} + \alpha_{zip.edu[i]} + \alpha_{eth.edu[i]} + \alpha_{eth.income[i]} + \alpha_{eth.age[i]},$$

where $y_i$ is the binary response variable (e.g., 1=Reactive for SARS-CoV2 antibodies, 0=non-reactive), male is a binary indicator for a participant $i$ reporting being male or female, $X_{ZIP}$ is the proportion of households that report being Hispanic in the ZIP code of participant $i$. Random intercepts, $\alpha$, were defined for each category of race/ethnicity (African American/Black; white; Hispanic; American Indian or Other; Asian or Pacific Islander; and two or more races), age in years (18-29, 30-44, 45-64, 65-74, 75+), education (bachelor's degree or not), income (2019 household income > $100,000 USD), household size (1, 2, 3, 4, 5, or ≥6 persons living in household), and 31 ZIP codes within study region as well as for interactions between ZIP code and race/ethnicity, ZIP code and education, race/ethnicity and education, race/ethnicity and income, and race/ethnicity and age were included (Table S-8). Based on the assumption that individuals of similar ages (e.g. those aged 18-29 and 30-44 vs. 65-74 and >75) may have similar behaviors and/or underlying infection risk, structured priors were used for age groups to allow for correlation between age groups.[10]

We allowed for spatial correlations at the ZIP code level, $\phi_{zip[i]}$,using the modified Besag-York-Mollié (BYM2) model.[11,12] The scale parameter for the BYM2 model was calculated with INLA::inla.scale.model in R using neighborhood graph data for our study region ZIP codes.[13] The BYM2 model is a reparameterization of the of the original Besag-York-Mollié (BYM) model which used random intercepts for non-spatial heterogeneity and Intrinsic Conditional Autoregressive (ICAR) components for spatial smoothing. The BYreM2 model replaces the random intercepts and ICAR components with a single parameter which is a weighted combination of the ICAR component, $\phi$ , and $\theta$, a parameter representing non-spatial heterogeneity:

$$\phi_j = \sigma\left(\phi_j \sqrt{\rho/s} + \theta\sqrt{1-\rho}\right)$$

$$\phi_j \vee \phi_{i\sim j} \sim N\left(\frac{\sum_{i\sim j}\phi_i}{d_i}, \frac{1}{d_i}\right)$$

$$\theta * \sim N\left(0, I_{N_j}\right)$$

$$\rho \sim U(0,1),\ \ \sigma \sim U(0,10),\ \ j = 31$$

#### 6.4 Race/ethnicity categories

Self-reported race and Hispanic ethnicity were collapsed into six categories: non-Hispanic White, non-Hispanic Black, Hispanic, Asian, and AMI/other. Participants who identified as either Mexican, Mexican American, Chicano, Puerto Rican, Cuban, or Other Hispanic were categorized as Hispanic. Participants who identified as South Asian, Chinese, Filipino, Japanese, Korean, Vietnamese, other Asian, Native Hawaiian, Guamanian or Chamorro, Samoan, or Other Pacific Islander were categorized as Asian or Pacific Islander. Participants who identified as American Indian, Alaskan Native, two or more, other, refuse, or don't know and non-Hispanic were categorized as American Indian or other.

#### 6.5 SARS-CoV-2 outcome prevalence across study region and within subgroups

We report population adjusted prevalence of our COVID-19 outcomes across the study region and within subgroups of our primary geographic and demographic variables of interest: ZIP code, sex, age, race/ethnicity, education, income, and household size. Posterior distributions of the prevalence outcomes for each poststratification strata were used to calculate the prevalence across the study region and within strata of interest. The mean of the posterior distribution of the poststratification estimates was used as the point estimate and 95% credible interval (95% CI) were the 2.5% and 97.5% quantiles of a posterior distribution. Density plots were used to verify that posterior distributions were unimodal.

#### 6.6 SARS-CoV-2 test-kit bias corrected seroprevalence

In addition to SARS-CoV-2 seroprevalence estimates, we also estimated cumulative SARS-CoV-2 seroprevalence at each study round adjusted for the net sensitivity and specificity of the SARS-CoV-2 antibody testing algorithm by incorporating an expression for the probability of testing seropositive as a function of net sensitivity and specificity of the testing algorithm and the seropositive prevalence in the population:[14]

$$p = \pi Se + (1 - \pi)(1 - Sp),$$

where p is the expected frequency of positive tests, $\pi$, is the population prevalence, $Se$ is the sensitivity of the assay, and $Sp$ is the specificity of the assay. Data on sensitivity and specificity for the antibody assays used these analyses are from validation studies reported by Wong et al (See Table S-5).[3]

##### *6.6.1 Net sensitivity and specific for assays within a study round*

In study rounds 2 and 3, serial testing was implemented to improve the sensitivity of the Ortho VITROS® Anti-SARS-CoV-2 Total Ig test (ORTHO). The follow expressions for the net sensitivity and specificity of a serial testing algorithm were used to combine sensitivity and specificity of each assay in the serial algorithm[15]:

$$Se_{net} = Se_1 Se_2$$

$$Sp_{net} = Sp_1 + Sp_2 - Sp_1 Sp_2,$$

where $Se_{net}$ and $Sp_{net}$ are the net sensitivity and specific of the serial testing algorithm, $Se_1$ and $Sp_1$ are the sensitivity and specificity of the first test, and $Se_2$ and $Sp_2$ are the sensitivity and specificity of the second test.

##### *6.6.2 Cumulative assay sensitivity and specificity*

Because we were interested in cumulative seroprevalence, defined as testing positive for antibodies to SARS-CoV-2 in the current or previous study rounds, we needed to account for the sensitivity and specificity in assays. For example, in Round 2, study participants were antibody positive if antibodies were detected in Round 1 and/or Round 2. In Round 3, study participants were antibody positive if antibodies were detected in Round 1, Round 2, and/or Round 3. Combining tests from two different rounds in this context is straight-forward under the criteria that if either test is positive, then the participant is positive for SARS-CoV-2 antibodies.[15] See table below for expressions for sensitivity and specificity for cumulative antibody positivity attributable to natural infection.

| Rounds tested | Sensitivity | Specificity |
|---|---|---|
| Round 1 only | $Se^{R1}$ | $Sp^{R1}$ |
| Round 2 only | $Se^{R2}$ | $Sp^{R2}$ |
| Round 3 only | $Se^{R3}$ | $Sp^{R3}$ |
| Rounds 1 and 2 | $Se^{R1} + Se^{R2} - (Se^{R1}Se^{R2})$ | $Sp^{R1}Sp^{R2}$ |
| Rounds 1, 2, and 3 | $Se^{R1} + Se^{R2} + Se^{R3} - (Se^{R1}Se^{R2}) - (Se^{R1}Se^{R3}) - (Se^{R2}Se^{R3}) + (Se^{R1}Se^{R2}Se^{R3})$ | $Sp^{R1}Sp^{R2}Sp^{R3}$ |

Abbreviations: Se, sensitivity; Sp, specificity; R, Round.

In Round 3, DBS samples that tested positive for antibodies to SARS-CoV-2 spike protein and negative to the SARS-CoV-2 nucleocapsid protein were considered to have antibodies from COVID-19 vaccination only. Therefore, a sample need to be positive for anti-spike antibodies using serial testing with Ortho S and ELISA S assays and negative for anti-nucleocapsid antibodies with Roche NC assay. The expressions for sensitivity and specificity for COVID-19 antibody positivity were as follows:

$$Se_{Spike} = Se_{Ortho}Se_{ELISA}$$

$$Sp_{spike} = Se_{Ortho} + Se_{ELISA} - Se_{Ortho}Se_{ELISA}$$

$$Se_{V} = Se_{Spike} + Se_{Roche} - Se_{Spike}Se_{Roche}$$

$$Sp_{V} = Sp_{spike}Sp_{Roche}$$

where $Se_{Spike}$ and $Sp_{spike}$ are the net sensitivity and specificity for the serial testing algorithm using the Ortho S and ELISA S assays, $Se_{Roche}$ and $Sp_{Roche}$ are the sensitivity and specificity of the Roche NC assay, and $Se_{V}$ and $Sp_{V}$ are the net sensitivity and specificity of testing positive for anti-spike antibodies and negative for anti-nucleocapsid antibodies.

See Table S-6 for estimates of net sensitivity and specificity from test-bias analyses implemented in NIMBLE.

#### 6.7 Mitigation analysis

The association between LCA cluster variable classes (high-risk vs. low-risk behavior) and seroprevalence and self-reported test positivity was estimated using the MRP model described above with random intercepts for the binary cluster variable levels and interactions between the cluster variable and ZIP code, age, race/ethnicity, education, and income (Table S-9). Prevalence differences (PD) and prevalence ratios (PR) were estimated by predicting the probability of the outcome given the LCA class and computing the difference or ratio,

$$PD = P(Y = 1|A = 1) - P(Y = 1|A = 0)$$

$$PR = P(Y = 1|A = 1) \;/\; P(Y = 1|A = 0),$$

where $Y$ is the binary antibody or self-test SARS-CoV-2 infection variable (1=SARS-CoV-2 positive, 0=SARS-CoV-2 negative) and $A$ is the binary LCA cluster variable (1="high-risk", 0="low-risk").

##### *6.7.1 Covariate imputation*

Missing covariate data was imputed with the median or mean value within a ZIP code if the missingness was less than <1% (sex, age, race/ethnicity, education, and household size). Data on income with missing in approximately 3-4% in the data depending on the study round. Binary household income (1: ≥\$100,000, 0: <\$100,000) was imputed within each nimble model using a logistic regression likelihood with sex, percent Hispanic within a ZIP code, and education as explanatory variables.

#### 6.8 Markov chain Monte Carlo (MCMC)

NIMBLE[16,17] was used to implement all MRP models using MCMC sampling. For adjusted prevalence outcomes we used 30,000 total iterations with 10,000 burn-in iterations across 10 chains, with a thinning interval of five. Seroprevalence test-bias models ran for 50,000 total iterations with 10,000 burn-in iterations across 10 chains, with a thinning interval of 10. Thinning was used due to memory constraints. We performed several MCMC diagnostics including estimation of effective sample size, visual inspection of trace and density plots for each parameter of interest, Gelman and Rubin's convergence diagnostic, and autocorrelation plots.

##### *6.8.1 Priors*

*Random intercepts and fixed effect priors*

The prior for a vector of random intercepts for a variable $V$ with $K$ categories was:

$$\alpha_{v[k]} \sim N(0, \sigma_{v[k]})$$

$$\sigma_{v[k]} \sim N_{[0,\infty)}(0, 0.5) \text{ for } k \text{ in } 1..K.$$

The prior for a fixed effect coefficient for a binary variable $V$ was $\beta_v \sim N(0, 0.5)$. The prior for a fixed effect coefficient for a continuous variable $V$ was $\beta_v \sim N(0, 0.5/\sigma_v)$. The structured prior[10] for the five-level categorical age variable was:

$$\alpha_j^{Age} | \alpha_{j-1}^{Age}, \ldots, \alpha_1^{Age}, \alpha^{Age} \sim N\left(\alpha_{j-1}^{Age}, \left(\sigma^{Age}\right)^2\right),$$

$$\text{for } j = 2, \ldots, 5$$

$$\sigma^{Age} \sim N_{(0,\infty)}(0,1),$$

$$\sum_{j=1}^{J} \alpha_j^{Age} = 0.$$

*Antibody test-bias priors*

We adapted our specifications for the sensitivity and specificity of the SARS-CoV-2 antibody assays from Carpenter and Gelman 2020.[14] The number of positive tests positive tests for assay $k$ was binomially distributed:

$$y_k \sim Binomial(p_k, n_k)$$

$$logit(p_k) \sim N(\mu_k, \sigma_k),$$

where $y_k$ is the number of positive tests in $n_k$ tests observed for assay $k$, $p_k$ is the probability of observing $y_k$positive tests in $n_k$ tests. $p_k$ has a normal prior, $\mu_k$ and $\sigma_k$ are not random variables, they are specified to match the sensitivity and specificity point estimate and confidence from validation studies performed by Wong et al (Table S-5).[3]

#### 6.9 Nimble Models and Diagnostics

Nimble model code, priors, and MCMC diagnostic results are available at https://github.com/adams-cam/ebcovid_prev.

### 7 Supplemental Tables

**Table S-1.** ZIP code of residence of participants at each round of the study compared to study region population.

| | PS Strata† % | Round 1 no. (%) | Round 2 no. (%) | Round 3 no. (%) |
|---|---|---|---|---|
| Invited to round, N | | 7166 | 6242 | 5506 |
| Participated in study round, N | | 5501 (76.8) | 5603 (89.8) | 4806 (87.3) |
| City | | | | |
| Albany | 2.3 | 245 (4.5) | 258 (4.6) | 229 (4.8) |
| Berkeley | 16.1 | 1446 (26.3) | 1535 (27.4) | 1314 (27.3) |
| El Cerrito | 3.1 | 226 (4.1) | 257 (4.6) | 226 (4.7) |
| El Sobrante | 3.2 | 113 (2.1) | 116 (2.1) | 107 (2.2) |
| Emeryville | 4.0 | 309 (5.6) | 289 (5.2) | 236 (4.9) |
| Hercules | 3.0 | 66 (1.2) | 69 (1.2) | 58 (1.2) |
| Oakland‡ | 48.8 | 2417 (43.9) | 2417 (43.1) | 2061 (42.9) |
| Pinole | 2.4 | 44 (0.8) | 48 (0.9) | 38 (0.8) |
| Richmond | 9.8 | 502 (9.1) | 496 (8.9) | 440 (9.2) |
| San Pablo | 7.4 | 133 (2.4) | 118 (2.1) | 97 (2) |

*PS, Poststratification; USD, United States Dollar.

†Population percentages from synthetic poststratification tables generated from American Community Survey (ACS) and Public Use Microdata Sample data. Each cell contains the percentage of population age >18. Gender is not available in ACS data.

‡Piedmont included in Oakland.

**Table S-2.** Distribution of COVID-19 probable case definition variables.

| | | Round 1 | Round 2 | Round 3 |
|---|---|---|---|---|
| N | | 5501 | 5603 | 4806 |
| COVID-like symptoms[a] | | | | |
| | No | 4574 (83.1) | 5092 (90.9) | 4143 (86.2) |
| | Yes | 898 (16.3) | 339 ( 6.1) | 571 (11.9) |
| | Not reported | 29 ( 0.5) | 172 ( 3.1) | 92 ( 1.9) |
| COVID close contact[a] | | | | |
| | No | 4720 (85.8) | 4873 (87.0) | 4370 (90.9) |
| | Yes | 101 ( 1.8) | 75 ( 1.3) | 57 ( 1.2) |
| | Not reported | 680 (12.4) | 655 (11.7) | 379 ( 7.9) |
| COVID probable case[a] | | | | |
| | No | 4800 (87.3) | 4938 (88.1) | 4419 (91.9) |
| | Yes | 21 ( 0.4) | 10 ( 0.2) | 8 ( 0.2) |
| | Not reported | 680 (12.4) | 655 (11.7) | 379 ( 7.9) |

Abbreviations: DBS, dried blood spot; QNS, Quantity not sufficient
[a]COVID-like symptoms, COVID close-contact, and COVID probable case were defined by Council and State Territorial epidemiologists.[4]

**Table S-3.** Self-reported mask wearing behavior during each study round

| | **Round 1** | **Round 2** | **Round 3** |
|---|---|---|---|
| Mask wearing behavior n (%) | | | |
| Ever wear a mask=Yes | 5432 (99.3) | 5420 (99.8) | 4694 (99.6) |
| Wear during leisure/exercise=Yes | 4433 (98.9) | 4619 (99.1) | 3951 (99.1) |
| Wear at work=Yes | 1259 (91.9) | 1488 (93.2) | 1251 (95.0) |
| Wear while shopping=Yes | 3824 (88.0) | 4041 (92.2) | 3621 (94.8) |
| Wear while home=Yes | 399 (7.3) | 556 (10.2) | 475 (10.1) |

**Table S-4.** Characteristics of study participants stratified by high-risk and low-risk mitigation behavior.

| | Round 1 | | | Round 2 | | | Round 3 | | |
|---|---|---|---|---|---|---|---|---|---|
| | **High Risk** | **Low Risk** | | **High Risk** | **Low Risk** | | **High Risk** | **Low Risk** | |
| N | 1599 | 3871 | *P* | 977 | 4452 | *P* | 1084 | 3630 | *P* |
| Sex | | | 0.999 | | | 0.001 | | | 0.089 |
| Female | 1018 (63.7) | 2465 (63.7) | | 573 (58.7) | 2874 (64.6) | | 718 (66.2) | 2299 (63.4) | |
| Male | 581 (36.3) | 1404 (36.3) | | 403 (41.3) | 1577 (35.4) | | 366 (33.8) | 1330 (36.6) | |
| Age, years | | | <0.001 | | | <0.001 | | | <0.001 |
| 18 – 29 | 170 (10.6) | 296 (7.6) | | 80 (8.2) | 281 (6.3) | | 67 (6.2) | 213 (5.9) | |
| 30 - 44 | 620 (38.8) | 1138 (29.4) | | 360 (36.9) | 1231 (27.7) | | 245 (22.6) | 1049 (28.9) | |
| 45 – 64 | 621 (38.9) | 1423 (36.8) | | 379 (38.8) | 1703 (38.3) | | 396 (36.6) | 1464 (40.4) | |
| 65 – 74 | 161 (10.1) | 764 (19.7) | | 131 (13.4) | 919 (20.7) | | 254 (23.5) | 713 (19.7) | |
| 75 + | 26 (1.6) | 249 (6.4) | | 26 (2.7) | 313 (7.0) | | 120 (11.1) | 188 (5.2) | |
| Race | | | <0.001 | | | <0.001 | | | <0.001 |
| African American/Black | 75 (4.7) | 192 (5.0) | | 21 (2.2) | 148 (3.3) | | 51 (4.7) | 87 (2.4) | |
| Native American/ Alaskan Native or Other | 32 (2.0) | 91 (2.4) | | 16 (1.6) | 89 (2.0) | | 22 (2.0) | 58 (1.6) | |
| Asian/Pacific Islander | 244 (15.3) | 609 (15.7) | | 160 (16.4) | 580 (13.0) | | 146 (13.5) | 513 (14.1) | |
| Hispanic | 312 (19.5) | 540 (14.0) | | 122 (12.5) | 480 (10.8) | | 129 (11.9) | 377 (10.4) | |
| Two or more races | 155 (9.7) | 345 (8.9) | | 92 (9.4) | 288 (6.5) | | 65 (6.0) | 263 (7.3) | |
| White | 781 (48.8) | 2092 (54.1) | | 565 (57.9) | 2862 (64.4) | | 668 (61.8) | 2329 (64.2) | |
| Education | | | <0.001 | | | 0.251 | | | <0.001 |
| College degree | 1392 (87.1) | 3519 (91.1) | | 901 (92.4) | 4058 (91.2) | | 956 (88.3) | 3368 (92.8) | |
| No college degree | 206 (12.9) | 345 (8.9) | | 74 (7.6) | 391 (8.8) | | 127 (11.7) | 260 (7.2) | |

**Table S-5.** Antibody assays used to detect antibodies against SARS-CoV-2 spike and nucleocapsid proteins.

| | **Round 1** | **Round 2** | **Round 3** | **Round 3** |
|---|---|---|---|---|
| Antibody Target | Spike | Spike | Spike | Nucleocapsid |
| Assay #1 | | | | |
| Name | VITROS® Anti-SARS-CoV-2 Total Ig | VITROS® Anti-SARS-CoV-2 Total Ig | VITROS® Anti-SARS-CoV-2 Total Ig | VITROS® Anti-SARS-CoV-2 Total Ig |
| DBS tested | All | All | All | All |
| Criteria | S/C ≥ 1: Reactive<br>S/C <1: Non-reactive | S/C ≥ 0.7: ELISA<br>S/C < 0.7: Non-reactive | S/C ≥ 0.7: ELISA<br>S/C < 0.7: Non-reactive | S/C ≥ 0.7: Roche<br>S/C < 0.7: Non-reactive |
| Se/Sp (95% CI) | Se: 80.6 (64.0 - 91.8)<br>Sp: 1 (63.1 - 1) | Se: 88.9 (73.9 - 96.9)<br>Sp: 1 (63.1 - 1) | Se: 88.9 (73.9 - 96.9)<br>Sp: 1 (63.1 - 1) | Se: 88.9 (73.9 - 96.9)<br>Sp: 1 (63.1 - 1) |
| Assay #2 | | | | |
| Name | | ELISA IgG | ELISA IgG | Roche NC Total Ig |
| DBS tested | | Ortho S/C ≥ 0.7 | Ortho S/C ≥ 0.7 | Ortho S/C ≥ 0.7 |
| Criteria | | OD ≥ 0.34: Reactive<br>OD < 0.34: Non-reactive | OD ≥ 0.34: Reactive<br>OD < 0.34: Non-reactive | S/C ≥ 0.0465: Reactive<br>S/C < 0.0465 Non-reactive |
| Se/Sp (95% CI) | | Se: 0.972 (88.7 - 99.9)<br>Sp: 1 (87.7 - 1) | Se: 0.972 (88.7 - 99.9)<br>Sp: 1 (87.7 - 1) | Se: 86.7 (69.3 - 96.2)<br>Sp: 97.9 (94.8 - 99.4) |

Se, sensitivity; Sp, specificity. Data on sensitivity and specificity from Wong et al.[18]

**Table S-6.** Net sensitivity and specificity estimated in NIMBLE MRP models for antibody testing algorithms using for population-adjusted antibody prevalence test bias analyses.

| Testing period | SARS-CoV-2 Outcome[a] | Rounds included for antibody testing[b] | Net Sensitivity % (95% CI)[c] | Net Specificity % (95% CI)[c] |
|---|---|---|---|---|
| Round 1 | SARS-COV-2 Natural Infection | R1 | 79.91 (66.01, 90.78) | 99.57 (99.3, 99.79) |
| Rounds 1-2 | SARS-COV-2 Natural Infection | R1 | 80.65 (67.23, 91.03) | 99.48 (99.16, 99.75) |
| | | R2 | 86.5 (75.63, 94.24) | 100 (99.98, 100) |
| | | Net R1+R2 | 97.38 (94.2, 99.18) | 99.48 (99.16, 99.75) |
| Rounds 1-3 | SARS-COV-2 Natural Infection | R1 | 80.54 (67.1, 91.06) | 99.2 (97.68, 99.82) |
| | | R2 | 86.37 (75.22, 94.24) | 100 (99.98, 100) |
| | | R3 | 76.84 (62.88, 88.1) | 99.98 (99.94, 100) |
| | | R1+R2 | 97.35 (94.15, 99.17) | 99.19 (97.67, 99.82) |
| | | R1+R2+R3 | 99.39 (98.48, 99.84) | 97.33 (94.14, 99.15) |
| Round 3 | COVID-19 Vaccination | R3 | 79.61 (68.26, 89.89) | 99.99 (99.99, 99.99) |

[a]SARS-CoV-2 outcomes for antibody test bias analyses were: 1) cumulative SARS-CoV-2 natural infection antibody positivity, defined as testing antibody positive in the current or previous round, and 2) COVID-19 vaccination antibody positivity, defined as testing positive for the SARS-CoV-2 spike protein and testing negative for the SARS-CoV-2 nucleocapsid protein in Round 3

[b]R1=Tested in Round 1 only, R2=Tested in Round 2 only, R3=Tested in Round 3 only, R1+R2=Tested in Rounds 1 and 2; R1+R2+R3=Tested in Rounds 1, 2, and 3.

[c]Net sensitivity and specificity were estimated within MRP models using NIMBLE. See Supplemental Methods section 6 for details.

**Table S-7: Survey questions concerning behaviors related to infection mitigation in rounds 1, 2, and 3.**

| Domain | Questions<br>(Response: Yes, No, Don't know, Refuse) |
|---|---|
| Masks | • In the past 7 DAYS have you worn a protective mask inside your home?<br>• In the past 7 DAYS have you worn a protective mask at work?<br>• In the past 7 DAYS have you worn a protective mask while travelling?<br>• In the past 7 DAYS have you worn a protective mask while shopping?<br>• In the pat 7 DAYS have you worn a protective mask while doing leisure activities outside? |
| Leaving home | • Thinking about the past 7 DAYS, did you leave your house?<br>• Thinking about the past 7 DAYS, did you leave your house for work?<br>• Thinking about the past 7 DAYS, did you leave your house for shopping?<br>• Thinking about the past 7 DAYS, did you leave your house for leisure/exercise?<br>• Thinking about the past 7 DAYS, did you leave your house for medical/healthcare?<br>• Thinking about the past 7 DAYS, did you leave your house to care for a relative?<br>• Thinking about the past 7 DAYS, did you leave your house for another reason? |
| Transportation | • Was your form of transportation when leaving your residence walking or biking?<br>• Was your form of transportation when leaving your residence public transportation?<br>• Was your form of transportation when leaving your residence a personal automobile or motorcycle?<br>• Was your form of transportation when leaving your residence a car service such as Uber, Lyft or Taxi?<br>• Was your form of transportation when leaving your residence another form? |
| Travel | Total count and yes/no within each of the following questions:<br>• Which country/countries have you traveled outside of the UNITED STATES in the PAST 2 WEEKS?<br>• Which country/countries have you traveled outside of the UNITED STATES since December 2019 or the last questionnaire and the 2 weeks PRIOR TO TODAY'S DATE?<br>• Which states have you traveled to outside California in the PAST 2 WEEKS?<br>• Which states have you traveled to outside California since December 2019 or the last questionnaire and the 2 weeks PRIOR TO TODAY'S DATE?<br>• Which California counties outside of your residence have you traveled to in the PAST 2 WEEKS?<br>• Which California counties outside of your residence have you traveled to since December 2019 or the last questionnaire and the 2 weeks PRIOR TO TODAY'S DATE? |
| Gathering attendance | • In the past 7 DAYS have you been to a gathering with a maximum of 10 people?<br>• In the past 7 DAYS have you been to a gathering with a maximum of 20 people?<br>• In the past 7 DAYS have you been to a gathering with a maximum of 50 people?<br>• In the past 7 DAYS have you been to a gathering with a maximum of 100 people?<br>• In the past 7 DAYS have you been to a gathering with more than 100 people?<br>• In the past 7 DAYS have you been to no gatherings? |
| Work exposures | • Have you had to continue to work even though you are in close contact with people who might be infected (e.g., customers, patients, co-workers)?<br>• Has someone in your household had to continue to work even though they are in close contact with people who might be infected (e.g., customers, patients, co-workers)?<br>• If you currently work in the healthcare field, do you come in contact with patients (within 6 ft) with confirmed COVID-19 infection? |

| | |
|---|---|
| | • Does anyone currently living in your household, not including yourself, work in the healthcare field? |

**Table S-8.** MRP logistic regression hierarchical model for SARS-CoV-2 outcomes.

| Variable | Description | Type | Number of groups |
|---|---|---|---|
| y | - Seroprevalence<br>- Self-report test positivity<br>- COVID-19 probable case | Binary Outcome variable | - |
| sex | Sex: Male or Female | Linear predictor | 2 |
| pct_span | Percent Spanish speaking within ZCTA | Linear predictor | - |
| age | Age | Varying intercept | 5 |
| race/eth | Race/ethnicity | Varying intercept | 6 |
| zip | BYM2 zip code variable | BYM2 varying intercept | 31 |
| edu | Education | Varying intercept | 2 |
| inc | Income | Varying intercept | 2 |
| hh | Household size | Varying intercept | 6 |
| zip_eth | Zip Code × ethnicity | Varying intercept | 31 × 6 = 186 |
| zip_edu | Zip Code × education | Varying intercept | 31 × 2 = 62 |
| eth_edu | Ethnicity × education | Varying intercept | 6 × 2 = 12 |
| eth_inc | Ethnicity × income | Varying intercept | 6 × 2 = 12 |
| eth_age | Ethnicity x age | Varying intercept | 6 x 5 = 30 |

**Table S-9.** MRP logistic regression hierarchical model for mitigation analysis.

| Variable | Description | Type | Number of groups |
|---|---|---|---|
| y | - Seroprevalence<br>- Self-report test positivity<br>- Covid-19 probable | Outcome variable | - |
| sex | Sex: Male or Female | Linear predictor | 2 |
| pct_span | Percent spanish speaking within ZCTA | Linear predictor | - |
| age | Age | Varying intercept | 5 |
| race/eth | Race/ethnicity | Varying intercept | 6 |
| zip | BYM2 zip code variable | BYM2 varying intercept | 31 |
| edu | Education | Varying intercept | 2 |
| inc | Income | Varying intercept | 2 |
| hh | Household size | Varying intercept | 6 |
| zip_eth | Zip Code × ethnicity | Varying intercept | 31 × 6 = 186 |
| zip_edu | Zip Code × education | Varying intercept | 31 × 2 = 62 |
| eth_edu | Ethnicity × education | Varying intercept | 6 × 2 = 12 |
| eth_inc | Ethnicity × income | Varying intercept | 6 × 2 = 12 |
| eth_age | Ethnicity x age | Varying intercept | 6 x 5 = 30 |
| exp | high vs. low-risk cluster variable | Varying intercept | 2 |
| exp_zip | high vs. low-risk cluster variable × Zip code | Varying intercept | 2 × 31 = 62 |
| exp_age | high vs. low-risk cluster variable × age | Varying intercept | 2 × 5 = 10 |
| exp_eth | high vs. low-risk cluster variable × race/eth | Varying intercept | 2 × 6 = 12 |
| exp_inc | high vs. low-risk cluster variable × income | Varying intercept | 2 × 2 = 4 |
| exp_edu | high vs. low-risk cluster variable × education | Varying intercept | 2 × 2 = 4 |

**Table S-10.** Population adjusted prevalence estimates of SARS-CoV-2 outcomes and respective 95% credible intervals across demographic and geographic strata *(see excel file.)*

### 8 Supplemental Figures

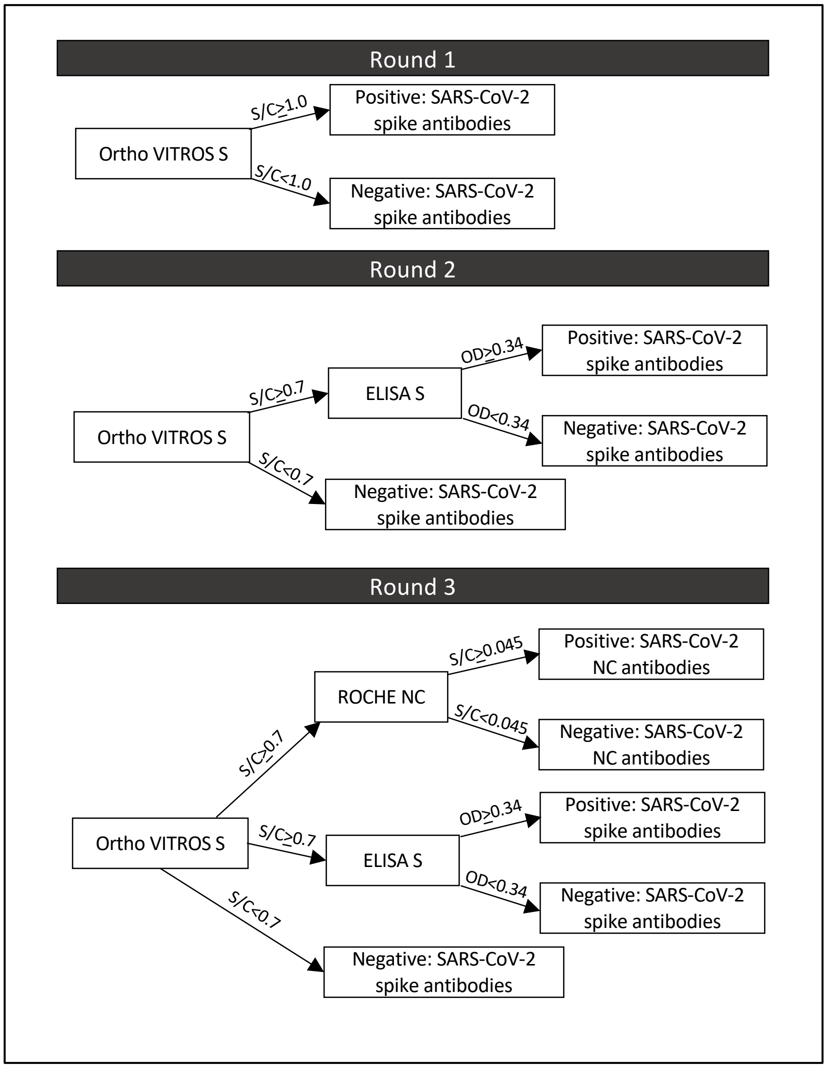

**Figure S-1.** Antibody testing algorithm for each study round. Antibodies against the spike (S) protein indicate prior or current SARS-CoV-2 infection or COVID-19 vaccination. Antibodies against the nucleocapsid (NC) protein indicate prior or current SARS-CoV-2 infection.

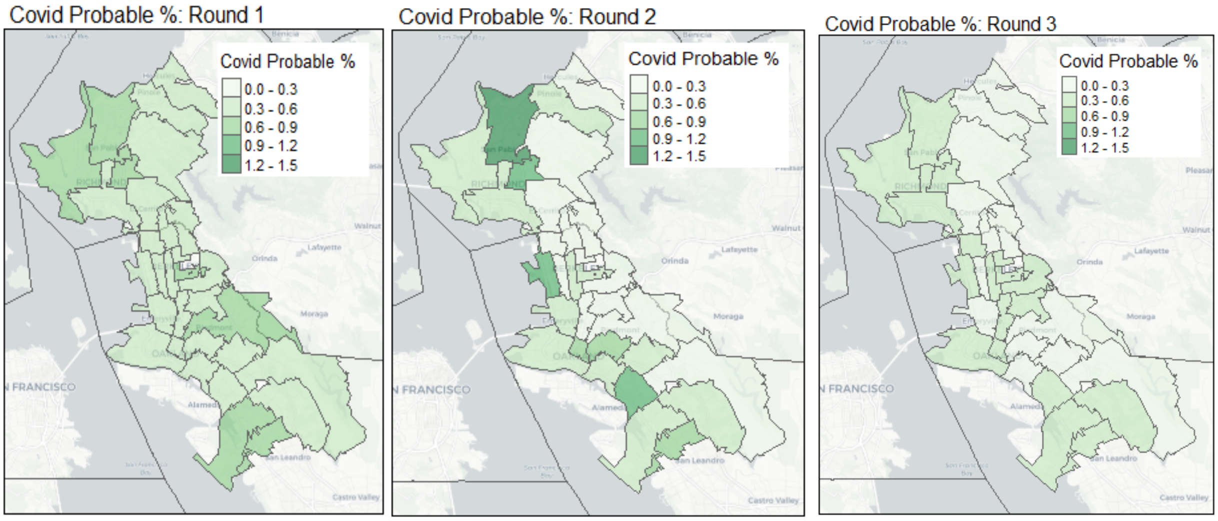

**Figure S-2.** Population-adjusted covid probable prevalence by zip code (July 2020-April 2021).

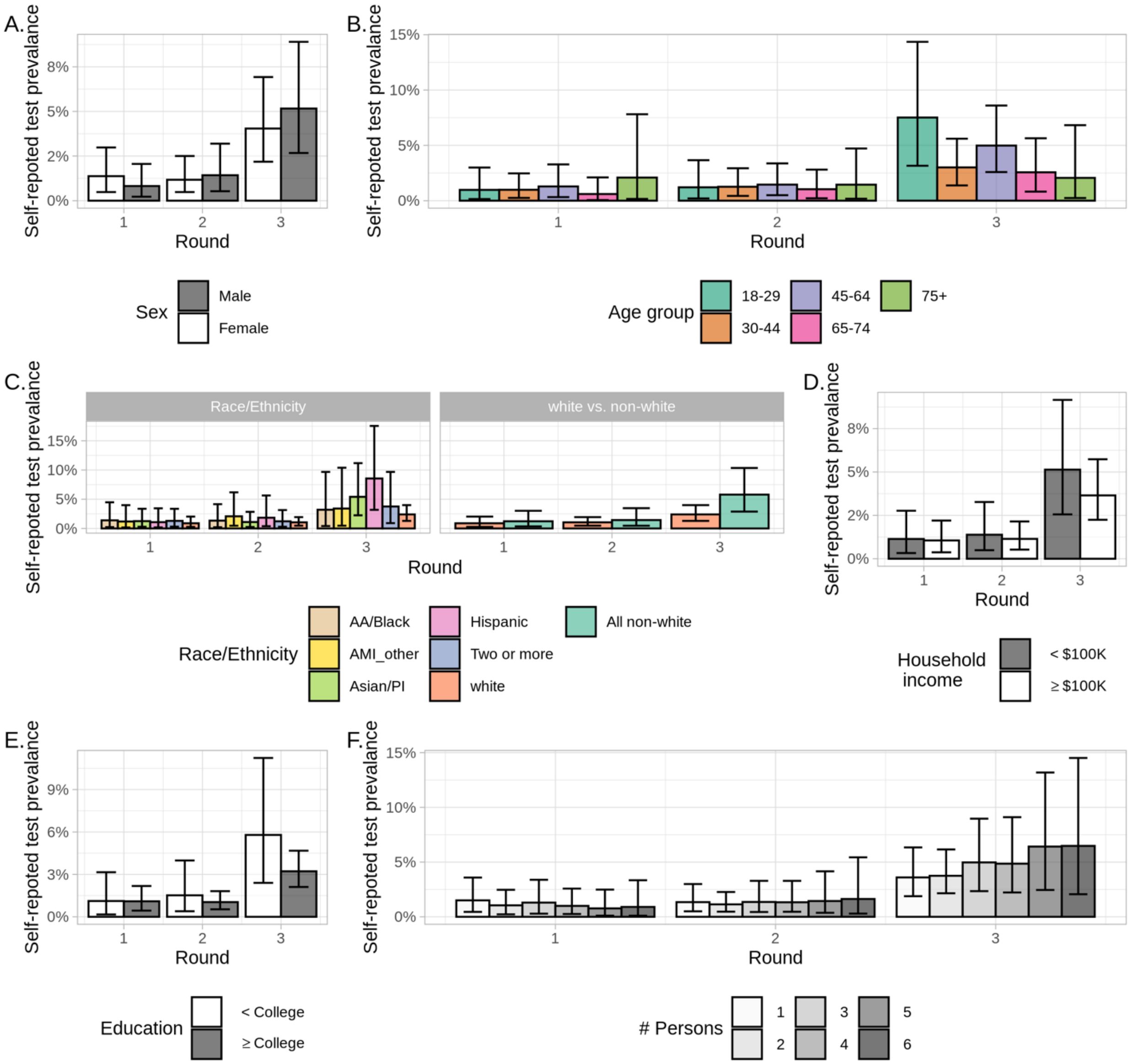

**Figure S-3.** Populated-adjusted self-reported COVID-19 test positivity in each study round among demographic subgroups, A) sex, B) age, C) race/ethnicity, D) income, E) education, and F) household size.

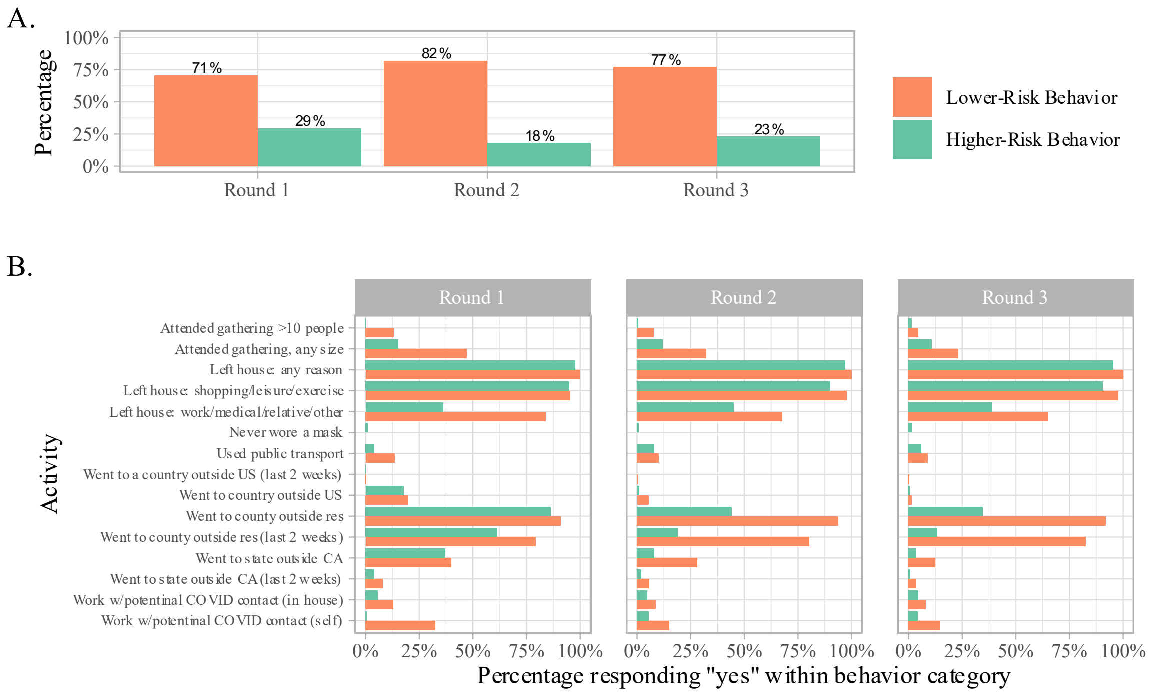

**Figure S-4.** Proportion of participants stratified by high- and low-risk mitigation behaviors in each study round, A) overall, and B) according to specific behavior.

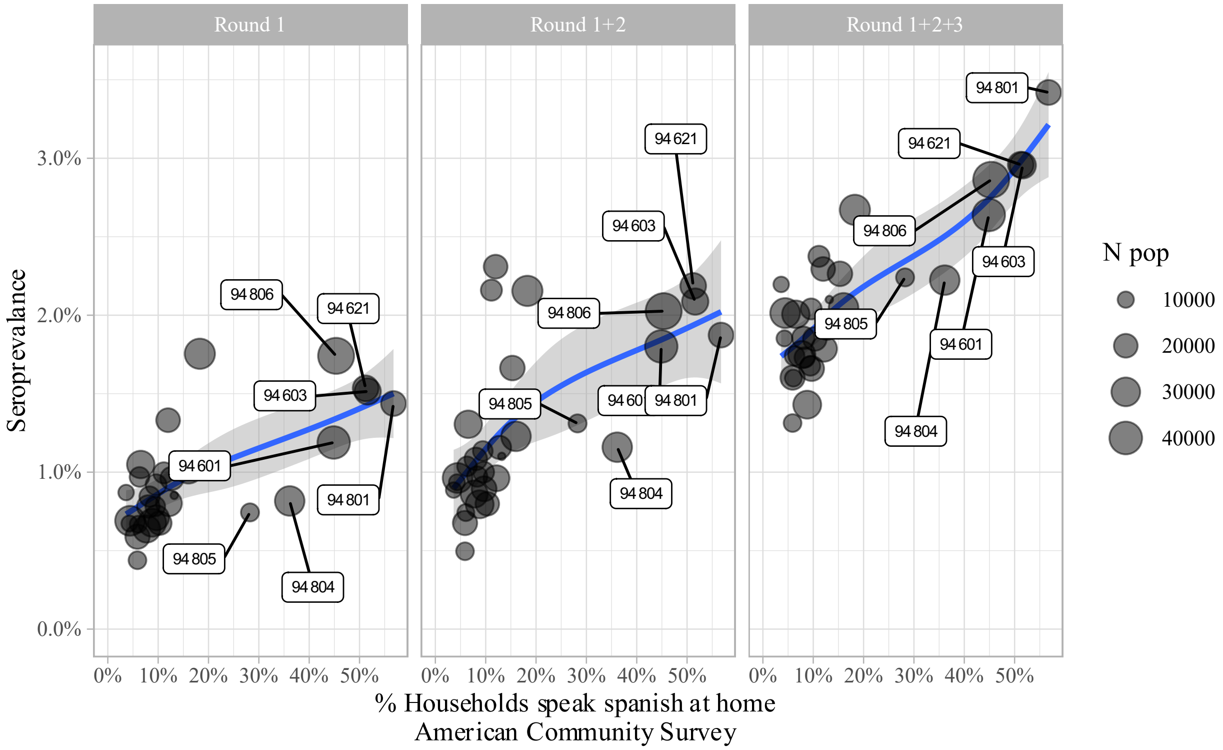

**Figure S-5.** Scatterplot of proportion of households who speak Spanish at home reported by the American Community Survey (x-axis) and cumulative population-adjusted SARS-CoV-2 seroprevalence (y-axis) within ZIP codes.

**A.**

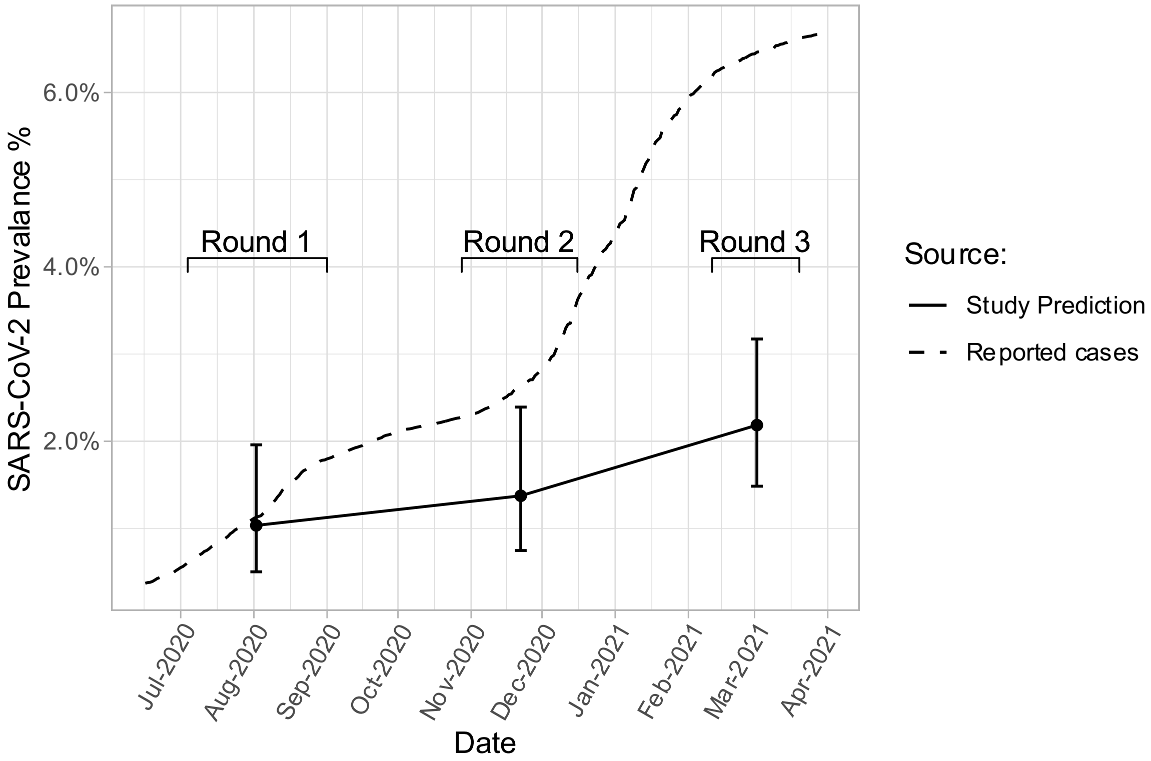

**Figure S-6.** Comparison of SARS-CoV-2 seroprevalence estimated in study to cumulative case prevalence reported by local public health agencies (July 2020-April 2021): A) Oakland/Piedmont, B) Albany, Berkeley, and Emeryville, and D) El Cerrito, Richmond, and San Pablo. The household response rate (RR) to the study invitation within each ZIP code is included in the upper left of each panel.

**B.**

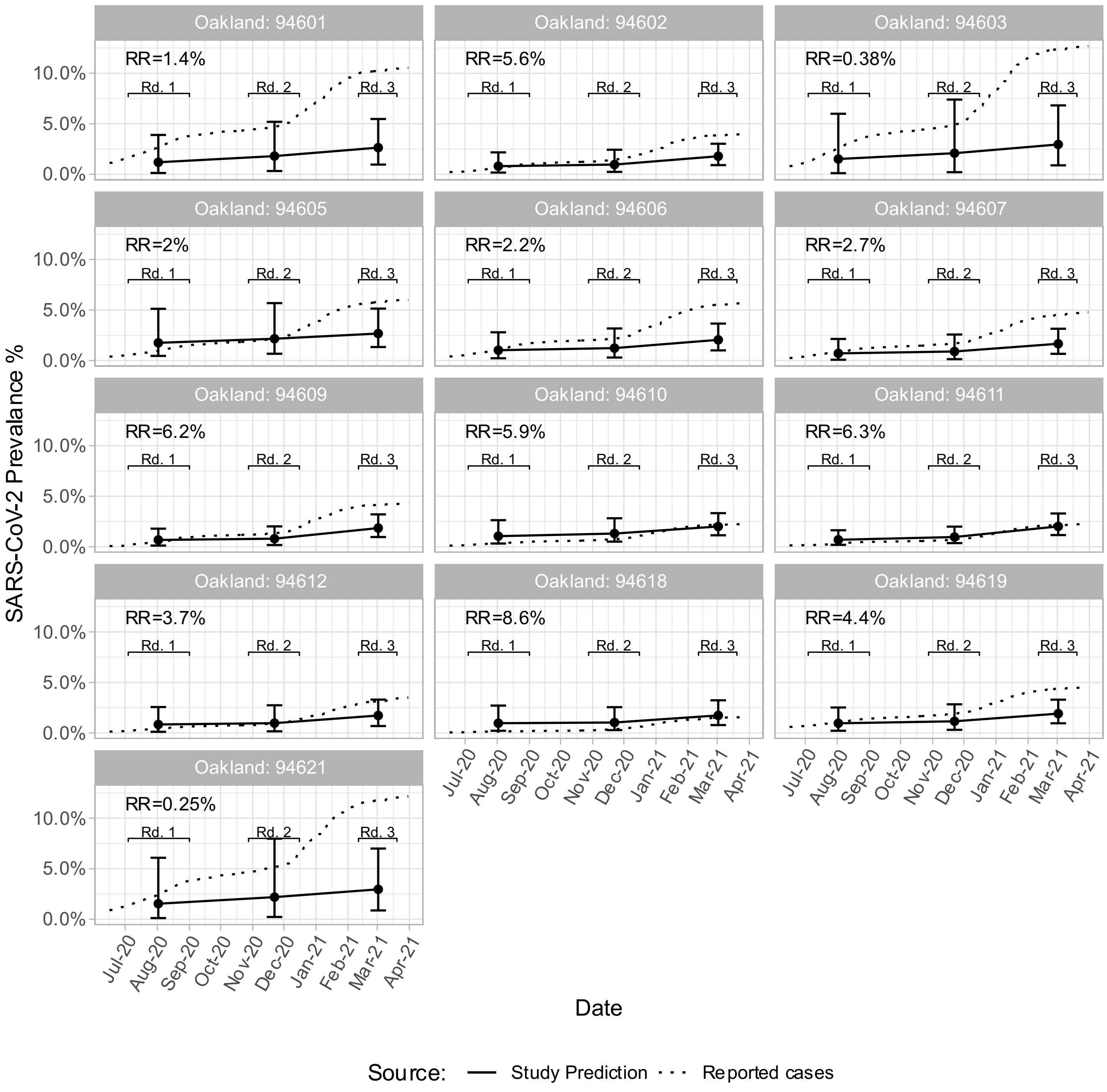

Oakland: 94601
RR=1.4%
Oakland: 94602
RR=5.6%
Oakland: 94603
RR=0.38%
Oakland: 94605
RR=2%
Oakland: 94606
RR=2.2%
Oakland: 94607
RR=2.7%
Oakland: 94609
RR=6.2%
Oakland: 94610
RR=5.9%
Oakland: 94611
RR=6.3%
Oakland: 94612
RR=3.7%
Oakland: 94618
RR=8.6%
Oakland: 94619
RR=4.4%
Oakland: 94621
RR=0.25%
Rd. 1
Rd. 2
Rd. 3
0.0%
5.0%
10.0%
SARS-CoV-2 Prevalance %
Jul-20
Aug-20
Sep-20
Oct-20
Nov-20
Dec-20
Jan-21
Feb-21
Mar-21
Apr-21
Date
Source:
Study Prediction
Reported cases

C.

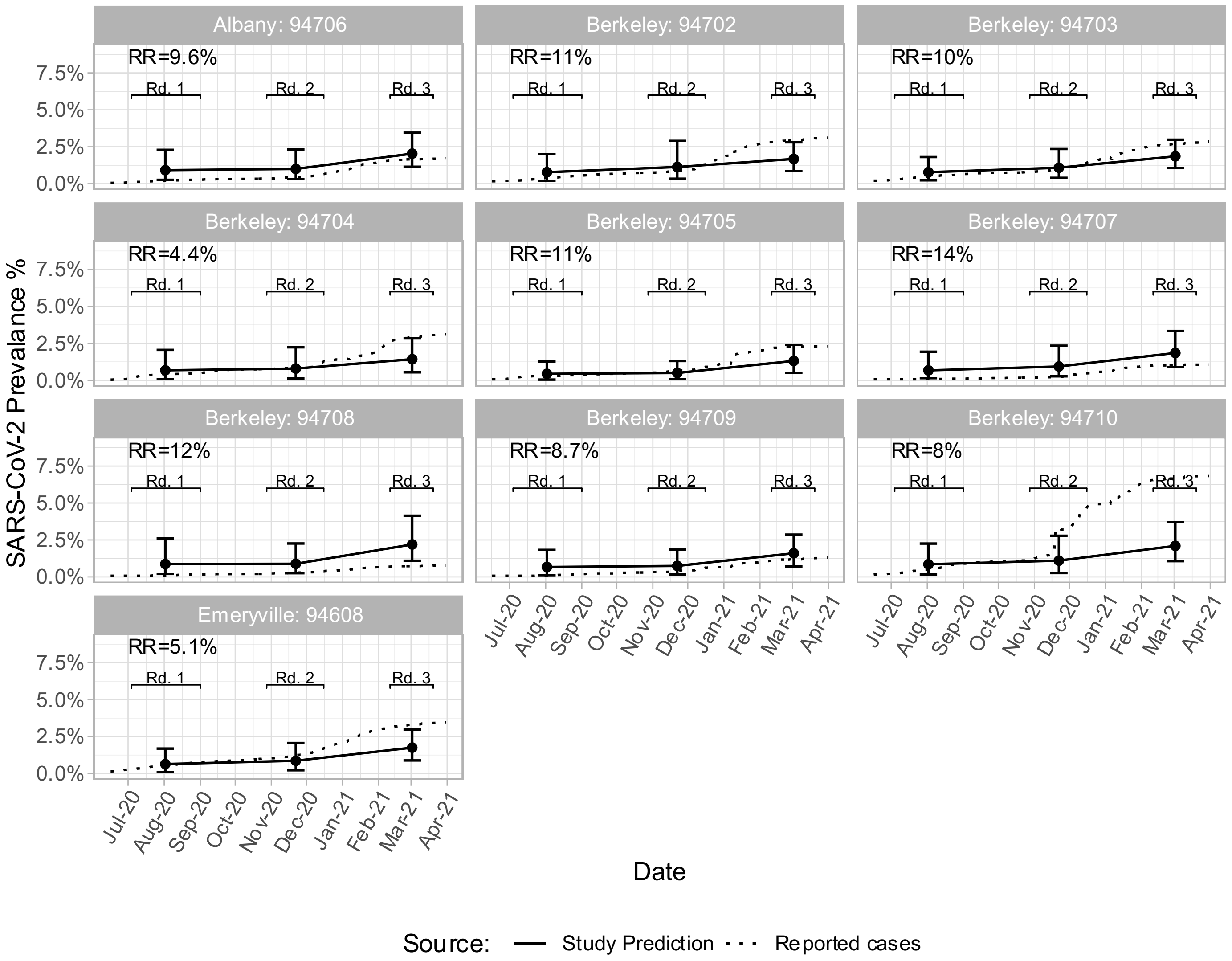
Albany: 94706
RR=9.6%
Berkeley: 94702
RR=11%
Berkeley: 94703
RR=10%
Berkeley: 94704
RR=4.4%
Berkeley: 94705
RR=11%
Berkeley: 94707
RR=14%
Berkeley: 94708
RR=12%
Berkeley: 94709
RR=8.7%
Berkeley: 94710
RR=8%
Emeryville: 94608
RR=5.1%
Rd. 1
Rd. 2
Rd. 3
0.0%
2.5%
5.0%
7.5%
SARS-CoV-2 Prevalance %
Jul-20
Aug-20
Sep-20
Oct-20
Nov-20
Dec-20
Jan-21
Feb-21
Mar-21
Apr-21
Date
Source:
Study Prediction
Reported cases

**D.**

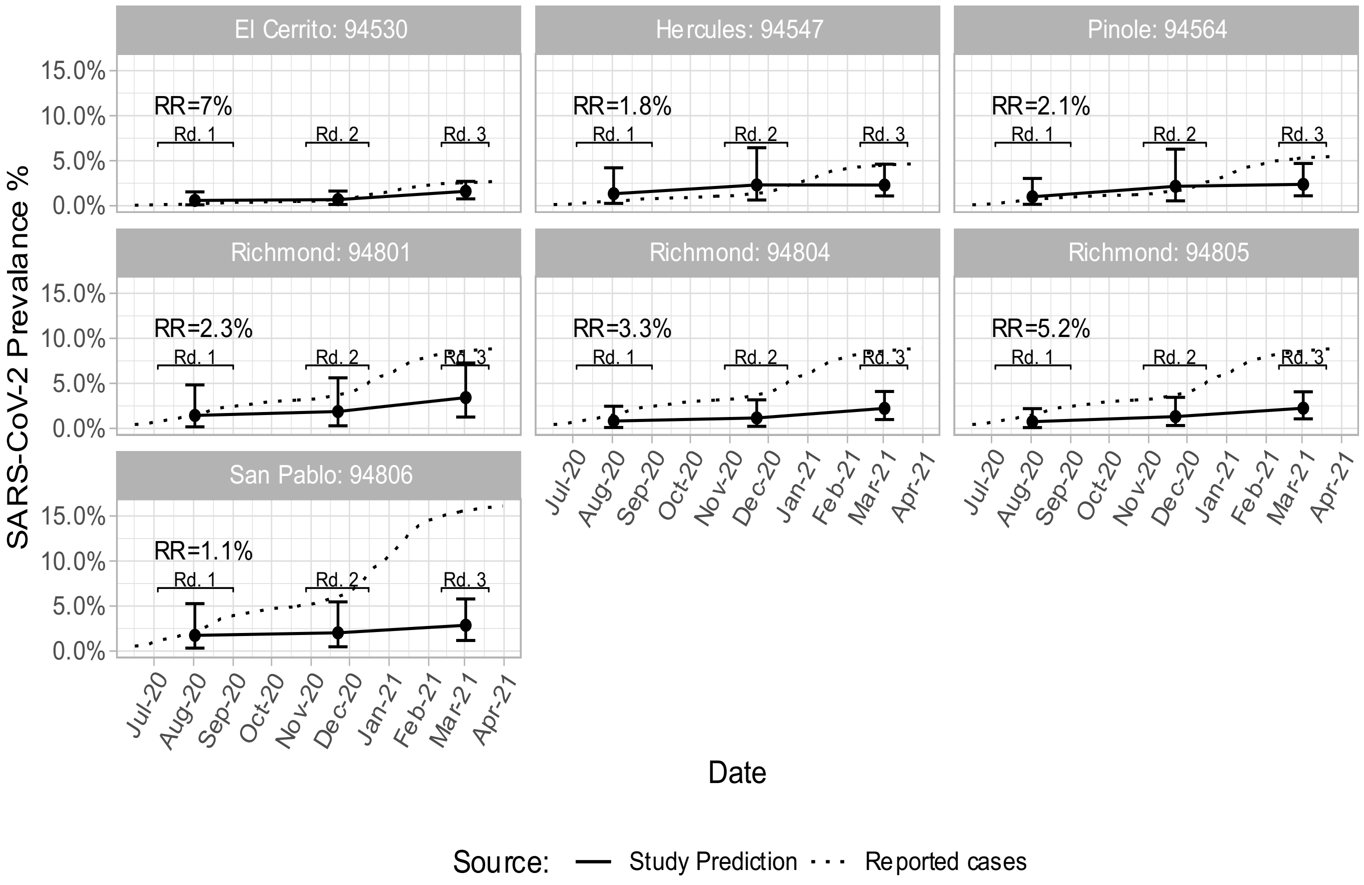

El Cerrito: 94530
Hercules: 94547
Pinole: 94564
Richmond: 94801
Richmond: 94804
Richmond: 94805
San Pablo: 94806
RR=7%
RR=1.8%
RR=2.1%
RR=2.3%
RR=3.3%
RR=5.2%
RR=1.1%
Rd. 1
Rd. 2
Rd. 3
15.0%
10.0%
5.0%
0.0%
SARS-CoV-2 Prevalance %
Jul-20
Aug-20
Sep-20
Oct-20
Nov-20
Dec-20
Jan-21
Feb-21
Mar-21
Apr-21
Date
Source:
Study Prediction
Reported cases

4 Council of State and Territorial Epidemiologists. Standardized surveillance case definition and national notification for 2019 novel coronavirus disease (COVID-19). 2020.

5 McLachlan, Jeffrey J, Thriyambakam Krishnan. The EM Algorithm and Extensions. New York: John Wiley & Sons, 1997.

6 Hagenaars JA, McCutcheon AL, editors. Applied Latent Class Analysis. Cambridge: Cambridge University Press, 2002 DOI:10.1017/CBO9780511499531.

7 Bandeen-Roche K, Miglioretti DL, Zeger SL, Rathouz PJ. Latent Variable Regression for Multiple Discrete Outcomes. *Journal of the American Statistical Association* 1997; **92**: 1375–86.

8 Gelman A, Little TC. Poststratification Into Many Categories Using Hierarchical Logistic Regression. *Survey Research* 1997; **23**: 127–35.

9 Leemann L, Wasserfallen F. Extending the Use and Prediction Precision of Subnational Public Opinion Estimation: EXTENDING USE AND PRECISION OF MrP. *American Journal of Political Science* 2017; **61**: 1003–22.

10 Gao Y, Kennedy L, Simpson D, Gelman A. Improving Multilevel Regression and Poststratification with Structured Priors. *Bayesian Analysis* 2021; **1**: 1–26.

11 Riebler A, Sørbye SH, Simpson D, Rue H. An intuitive Bayesian spatial model for disease mapping that accounts for scaling. *arXiv:160101180 [stat]* 2016; published online Jan 6. http://arxiv.org/abs/1601.01180 (accessed Sept 15, 2021).

12 Simpson D, Rue H, Riebler A, Martins TG, Sørbye SH. Penalising Model Component Complexity: A Principled, Practical Approach to Constructing Priors. *Statist Sci* 2017; **32**. DOI:10.1214/16-STS576.

13 van Niekerk J, Bakka H, Rue H, Schenk O. New frontiers in Bayesian modeling using the INLA package in R. *arXiv:190710426 [stat]* 2019; published online July 25. http://arxiv.org/abs/1907.10426 (accessed Sept 15, 2021).

14 Gelman A, Carpenter B. Bayesian analysis of tests with unknown specificity and sensitivity *. *medRxiv* 2020; : 2020.05.22.20108944-2020.05.22.20108944.

15 Weinstein S, Obuchowski NA, Lieber ML. Clinical Evaluation of Diagnostic Tests. *American Journal of Roentgenology* 2005; **184**: 14–9.

16 de Valpine P, Turek D, Paciorek CJ, Anderson-Bergman C, Lang DT, Bodik R. Programming with models: writing statistical algorithms for general model structures with NIMBLE. *Journal of Computational and Graphical Statistics* 2017; **26**: 403–13.

17 NIMBLE: MCMC, Particle Filtering, and Programmable Hierarchical Modeling. DOI:doi: 10.5281/zenodo.1211190.

18 Wong MP, Meas MA, Adams C, *et al.* Development and Implementation of Dried Blood Spot-based COVID-19 Serological Assays for Epidemiologic Studies. *medRxiv* 2021; : 2021.11.25.21266786.
